# A phenome-wide association study of methylated GC-rich repeats identifies a GCC repeat expansion in *AFF3* as a significant cause of intellectual disability

**DOI:** 10.1101/2023.05.03.23289461

**Authors:** Bharati Jadhav, Paras Garg, Joke J. F. A. van Vugt, Kristina Ibanez, Delia Gagliardi, William Lee, Mariya Shadrina, Tom Mokveld, Egor Dolzhenko, Alejandro Martin-Trujillo, Scott L. Gies, Clarissa Rocca, Mafalda Barbosa, Miten Jain, Nayana Lahiri, Katherine Lachlan, Henry Houlden, Benedict Paten, Genomics England Research Consortium, Project MinE ALS Sequencing Consortium, Jan Veldink, Arianna Tucci, Andrew J. Sharp

## Abstract

GC-rich tandem repeat expansions (TREs) are often associated with DNA methylation, gene silencing and folate-sensitive fragile sites and underlie several congenital and late-onset disorders. Through a combination of DNA methylation profiling and tandem repeat genotyping, we identified 24 methylated TREs and investigated their effects on human traits using PheWAS in 168,641 individuals from the UK Biobank, identifying 156 significant TRE:trait associations involving 17 different TREs. Of these, a GCC expansion in the promoter of *AFF3* was linked with a 2.4-fold reduced probability of completing secondary education, an effect size comparable to several recurrent pathogenic microdeletions. In a cohort of 6,371 probands with neurodevelopmental problems of suspected genetic etiology, we observed a significant enrichment of *AFF3* expansions compared to controls. With a population prevalence that is at least 5-fold higher than the TRE that causes fragile X syndrome, *AFF3* expansions represent a significant cause of neurodevelopmental delay.

## INTRODUCTION

Tandem repeat expansions (TREs) are a class of recurrent mutation in which a tandemly repeated DNA motif, *e.g.*, poly(CGG), undergoes expansion of repeat number to unusually large size. Expansions of tandem repeats (TRs) were first recognized as a mechanism of genetic disease in the early 1990s^1–3^ and, since this initial discovery, more than 50 inherited disorders, almost all of which are dominant in nature, are now known to be caused by TREs^4^. One class of TRE that has been linked with a variety of both congenital and late-onset disorders are composed of GC-rich TR motifs typically located within gene promoters or 5’UTRs^5^. The best exemplar of these is the CGG TRE within the 5’UTR of *FMR1* that causes fragile X syndrome, one of the most common genetic causes of intellectual disability and autism. Although this repeat is highly polymorphic in the normal population, in individuals with fragile X syndrome this TR expands to >200 copies, becoming highly methylated, accompanied by epigenetic silencing of *FMR1* and the presence of a cytogenetically-visible chromosomal fragile site at Xq28 after which the syndrome is named^6^. In addition, intermediate between common polymorphic length variation of the TR and the much rarer, methylated “full mutation” expansions that cause fragile X syndrome, unmethylated moderate-sized expansions (55-200 copies) of the *FMR1* CGG TR occur in relatives of probands with fragile X syndrome. Termed “premutations” due to their propensity to further expand upon meiotic transmission, these shorter expanded alleles drive increased expression of *FMR1* and are now recognized as a cause of two other disorders, namely fragile X premature ovarian insufficiency (FXPOI) and fragile X tremor ataxia syndrome (FXTAS), the latter of which occurs in up to 80% of male carriers^7,8^. Since the discovery of the molecular basis of *FMR1*-related disorders, other similar promoter-associated GC-rich TREs have been implicated as causal variants underlying a variety of conditions, including epilepsy, oculopharyngodistal myopathy and neurodegenerative disorders^9–18^, raising the possibility that the human genome may harbor additional similar TREs that contribute to a range of pathologies.

Previously, we showed that the use of DNA methylation profiling represents a powerful method to identify rare methylated TREs^19^. Here, we applied this approach to analyze DNA methylation profiles from 5,750 individuals for whom PCR-free Illumina genome sequencing (GS) data were available, identifying 24 GC-rich promoter-associated TRs that become methylated when expanded. Given that expansions of similar GC-rich repeats have previously been implicated in multiple human disorders^4,5^, we hypothesized that these TREs represent strong candidates for modifiers of human disease risk. To investigate this, we used GS data to directly genotype these loci and performed a phenome-wide association study for TREs in 168,641 individuals from the UK Biobank (UKB), identifying numerous associations with human traits. Based on our findings, we further investigated the role of a TRE within *AFF3* as a contributor to neurodevelopmental disorders in the UK 100,000 Genomes cohort, confirming this as a clinically significant cause of intellectual disability.

## RESULTS

### Identification of repeat expansions associated with local hypermethylation

Profiling the methylomes of 5,750 samples, we identified a total of 2,124 differentially methylated regions (DMRs) (Supplementary Tables 1 and 2). Using ExpansionHunter Denovo to analyze available GS data, we performed genome-wide screening for TREs in these individuals, identifying 34 candidate TREs that overlapped hypermethylated DMRs. We then utilized ExpansionHunter to genotype each candidate TR that overlapped a hypermethylated DMR. Where expanded repeats exceed the sequencing read length, ExpansionHunter generates estimated allele sizes based on indirect metrics, such as the number of in-repeat reads. For GC-rich TREs that often form strong secondary structures, this can lead to uncertainty in the estimated genotypes and difficulty in distinguishing between unmethylated premutation alleles and methylated full mutations^20–21^. Despite this limitation, we observed many TR loci where individuals with local hypermethylation tended to have the longest TR alleles at that locus in the entire cohort. Using permutation analysis, 24 of the 34 loci tested showed a significant co-occurrence of hypermethylation and unusually long TR alleles in the same individuals (Figure 1), thus providing strong statistical evidence that the observed hypermethylation at each of these loci resulted from expansion of the underlying TR (Supplementary Table 3). At some of these 24 loci, a few individuals with hypermethylation were genotyped by ExpansionHunter as having two TR alleles within the normal range (Figure 1). We postulate that these may represent either a failure of ExpansionHunter to identify TREs (false negatives), or alternatively hypermethylation of the locus for reasons unrelated to the TR. In support of the former hypothesis, examination of read alignments^22^ in these individuals showed a small number of reads composed of long TR tracts, providing suggestive evidence of TREs that were missed by ExpansionHunter (Supplementary Figure 1). At the remaining 10 of 34 loci tested, permutation testing showed no association between the presence of hypermethylation and long TR alleles and these were excluded from further analysis.

**Figure 1.**
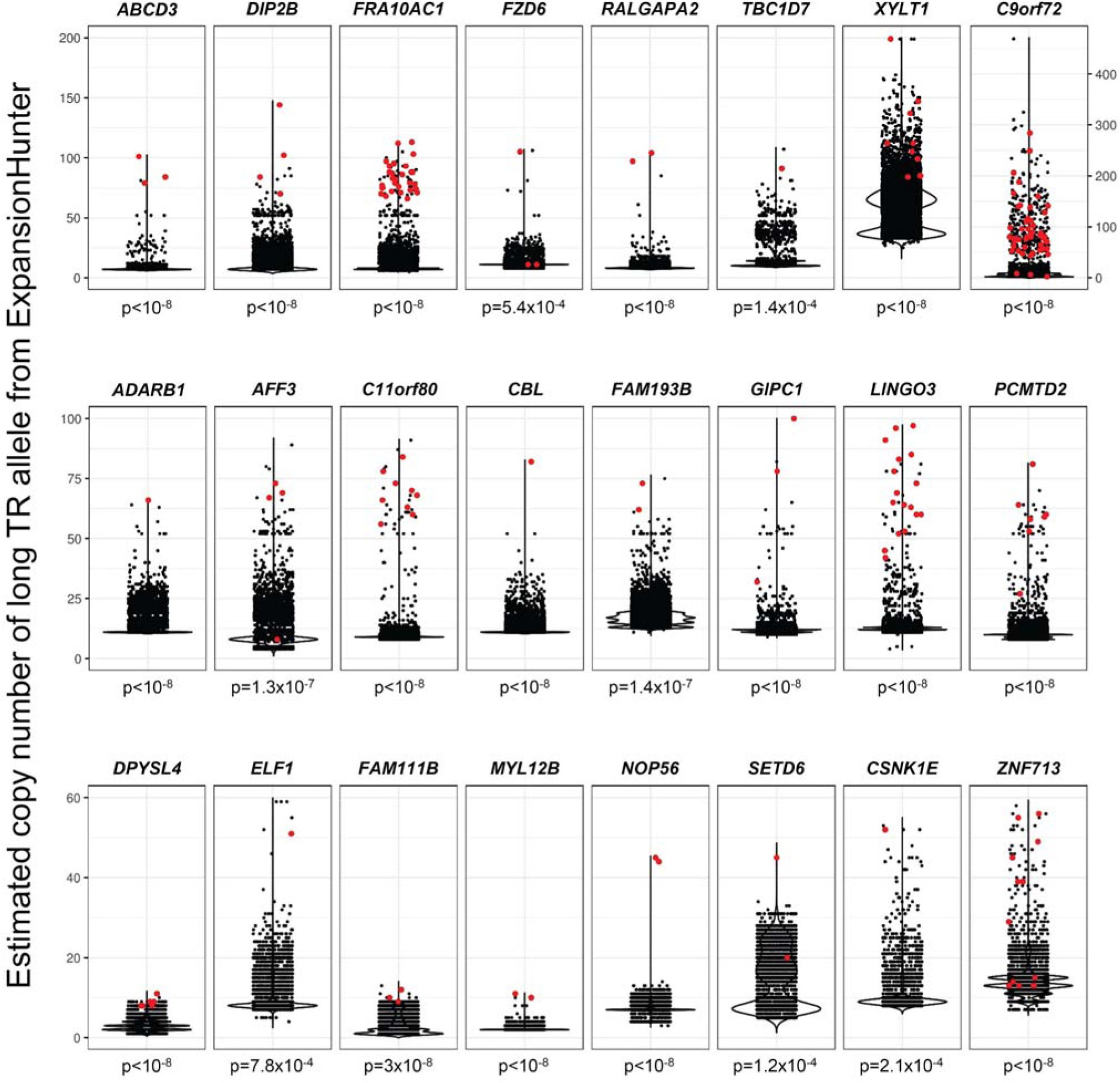
Confirmation of expanded GC-rich tandem repeats as the underlying cause of rare gains of methylation at 24 loci in the human genome. Each plot shows the copy number of the long TR allele in each sample derived from ExpansionHunter analysis of Illumina GS data. Individuals with hypermethylation of the locus are shown in red, individuals without hypermethylation are shown in black. P-values below each plot show the probability that the observed distribution of individuals with hypermethylation would occur by chance, derived from 100 million permutations. Full details of each TR locus are shown in Supplementary Table 3.

As expected, 20 of the 24 loci identified have previously been reported or suspected to undergo hypermethylation when expanded^13,14,16,19,23–30^ and seven have been associated with human disease^13–16,24,25,31,32^. However, we also identified methylated TREs that have not been previously described (Supplementary Table 3). Of note, we observed methylated GGC expansions within the 5’UTR of *GIPC1*, a locus where unmethylated and, thus presumably shorter TREs have recently been reported to cause oculopharyngodistal myopathy 2 (OPDM2)^15^ and methylated expansions of a GGCCTG repeat intronic within *NOP56* that cause SCA36^31^, thus implicating epigenetic effects of this TRE. While the majority of these methylated TREs were triplet repeats, a few were composed of longer TR motifs, including two 16mers.

### Confirmation of repeat expansions by long-read sequencing

We generated long-read GS to confirm the presence of three previously unreported TREs in samples that showed long TR alleles as genotyped by ExpansionHunter. PacBio GS confirmed expansions of a 16mer located in the 5’ UTR of *DPYSL4* (chr10:132,186,843-132,186,915), which consisted of 51-55 copies of the consensus motif CCGGGGGCGGGGCCTG and an expansion located within the 5’ UTR of *FAM193B* (chr5:177,554,490-177,554,531) consisting of 1,066-1,436 copies of a CCG repeat.

Using nanopore long-read GS, a single spanning read confirmed a TRE consisting of 473 copies of a GCC repeat located in the bidirectional promoter/5’ UTR region of *WBP4* and *ELF1*. Multiple other reads from the expanded allele produced very low quality sequence upon traversing the TR, which appeared to occur in a unidirectional fashion, a phenomenon we also observed with PacBio GS at *DPYSL4* and previously at other methylated GC-rich TREs that presumably have strong secondary structures that inhibit sequencing^19^. Despite this, we performed phasing of nanopore reads based on presence/absence of the TRE-containing haplotype and then analyzed the signal profiles, demonstrating that the expanded TR allele was highly methylated, while the normal TR allele was largely unmethylated (Figure 2).

**Figure 2.**
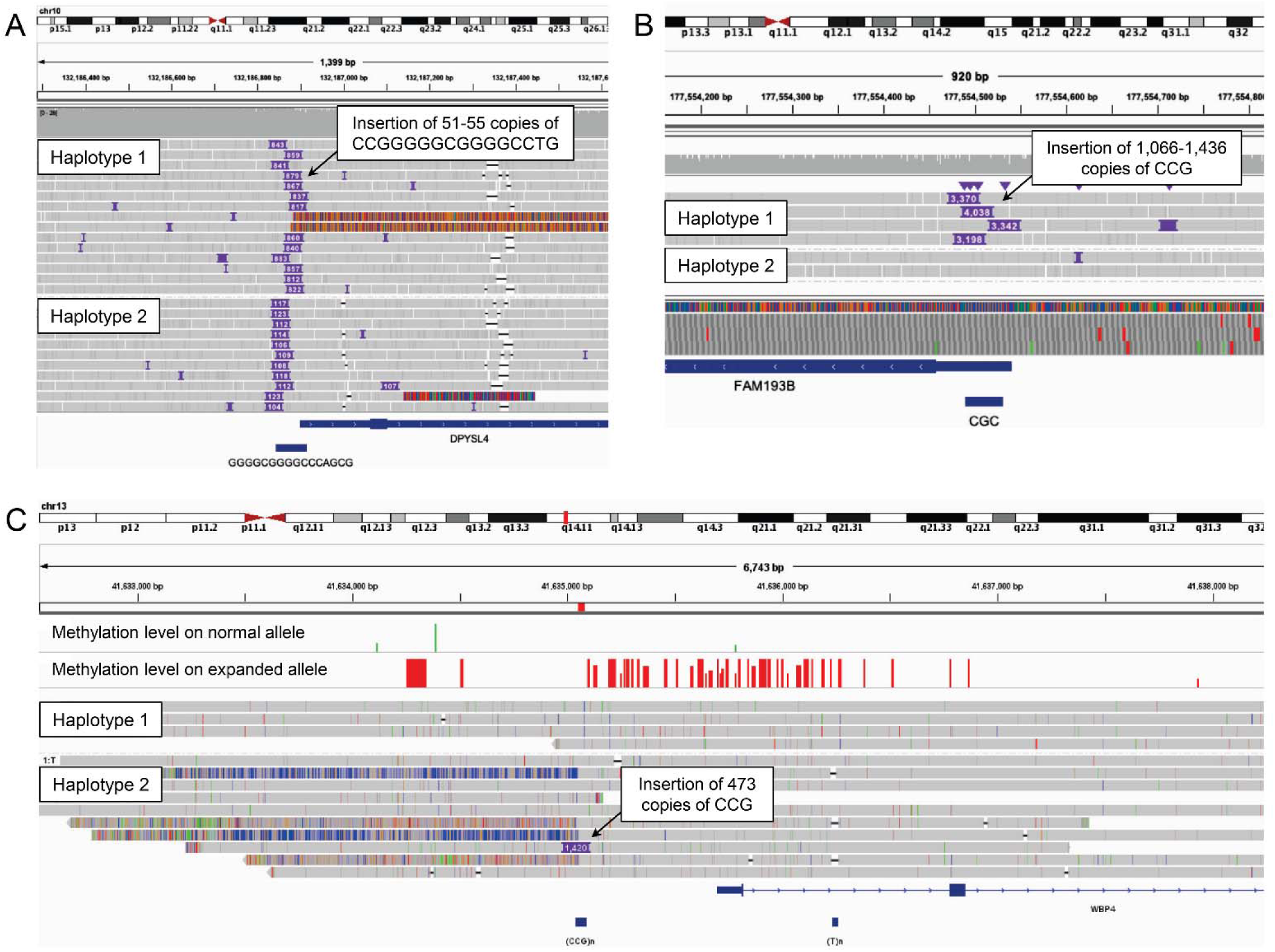
Validation of GC-rich repeat expansions using long-read sequencing. Screenshots from IGV showing PacBio GS of **(A)** A 16mer TRE within the 5’ UTR of *DPYSL4*, **(B)** A CGG TRE within the 5’ UTR of *FAM193B*. **(C)** Oxford Nanopore Technology GS of a CGG TRE in the bidirectional promoter/5’ UTR of *WBP4* / *ELF1*. In each case, purple boxes show the location and size of insertions in base pairs compared to hg38. By analyzing the signal profiles of the reads after phasing, we demonstrated that the expanded allele was highly methylated, while the normal TR allele was largely unmethylated. Images show read alignments in IGV. Below each track are shown gene and simple repeat annotations.

Notably, although ExpansionHunter correctly identified individuals carrying unusually long TR alleles at all three of these loci, the estimated TR lengths provided by ExpansionHunter were considerably shorter than the alleles observed directly with spanning long reads.

### Gene expression analysis in the GTEx cohort

To assess the functional effects of methylated TREs on gene expression, we utilized DNA methylation and RNAseq data generated by the GTEx project^33,34^. We identified 10 individuals with hypermethylation at *ADARB1*, *C11orf80*, *CSNK1E*, *FRA10AC1*, *PCMTD2* and *ZNF713*, indicating that they likely carry these TREs (Supplementary Figure 2). These individuals showed highly biased allelic expression (Supplementary Figure 3) and, in all but one case, this was accompanied by unusually low expression of the associated gene, thus indicating that these TREs result in allelic silencing *in cis*. The sole exception was an individual with moderately increased methylation of *ZNF713* who exhibited unusually high expression of this gene across nearly all tissues (Supplementary Figure 4).

### Phenome-wide association study of repeat expansions in the UK Biobank

As multiple GC-rich TREs are known to underlie a variety of human disorders^1,9–18^, we next sought to identify traits associated with the presence of GC-rich TREs through phenome-wide association studies (PheWAS). We used ExpansionHunter to analyze Illumina GS data from 168,641 unrelated individuals of European ancestry from the UKB, generating genotypes for 28 TRs, representing the 24 TREs associated with hypermethylation identified here in addition to four other GC-rich TRs from the literature where rare methylated TREs have been reported (Supplementary Table 3). In order to optimize our PheWAS pipeline to detect associations with expanded repeat alleles where the pathogenic threshold might vary widely among different loci, we performed outlier analysis using six different stringencies at each TR locus (Supplementary Table 4, Supplementary Figure 5) to identify sets of unusually long TR alleles representing putative TREs (Supplementary Table 5) and performed association analysis of expanded repeat alleles with 10,615 traits.

We identified 156 significant unique TRE:trait pairwise associations involving 17 different TREs at a threshold of 10% FDR (Supplementary Tables 6 and 7), indicating diverse negative effects of these TREs on human health. The most significant and numerous of these associations (n=93) occurred with expansions of the GGGGCC repeat within *C9orf72* that is known as a major cause of amyotrophic lateral sclerosis (ALS)^13,14^. We identified 350 individuals with a *C9orf72* TR allele >30 copies, indicating a prevalence of potentially pathogenic *C9orf72* TREs of 1 per 482 individuals (0.21%), in agreement with two prior studies that reported frequencies of 0.15%^35^ and 0.19%^21^. Based on the frequency of this mutation in ALS^36^, this carrier frequency is significantly above expectation and suggests pleiotropy and/or reduced penetrance, similar to a recent finding in Kennedy’s disease^37^. Although UKB data does not include a specific phecode for ALS, all of the observed traits associated with *C9orf72* expansions were consistent with the known symptoms, effects and treatments of ALS, including multiple measures of reduced brain volume, increased frequencies of depression, dysphagia, urinary tract infections, imaging studies of the central nervous system and higher death rates attributed to motor neuron disease and respiratory failure^38^ (Figure 3, Supplementary Table 6). We did not observe any evidence of variable penetrance in relation to size of the *C9orf72* expansion (Supplementary Figure 6).

**Figure 3.**
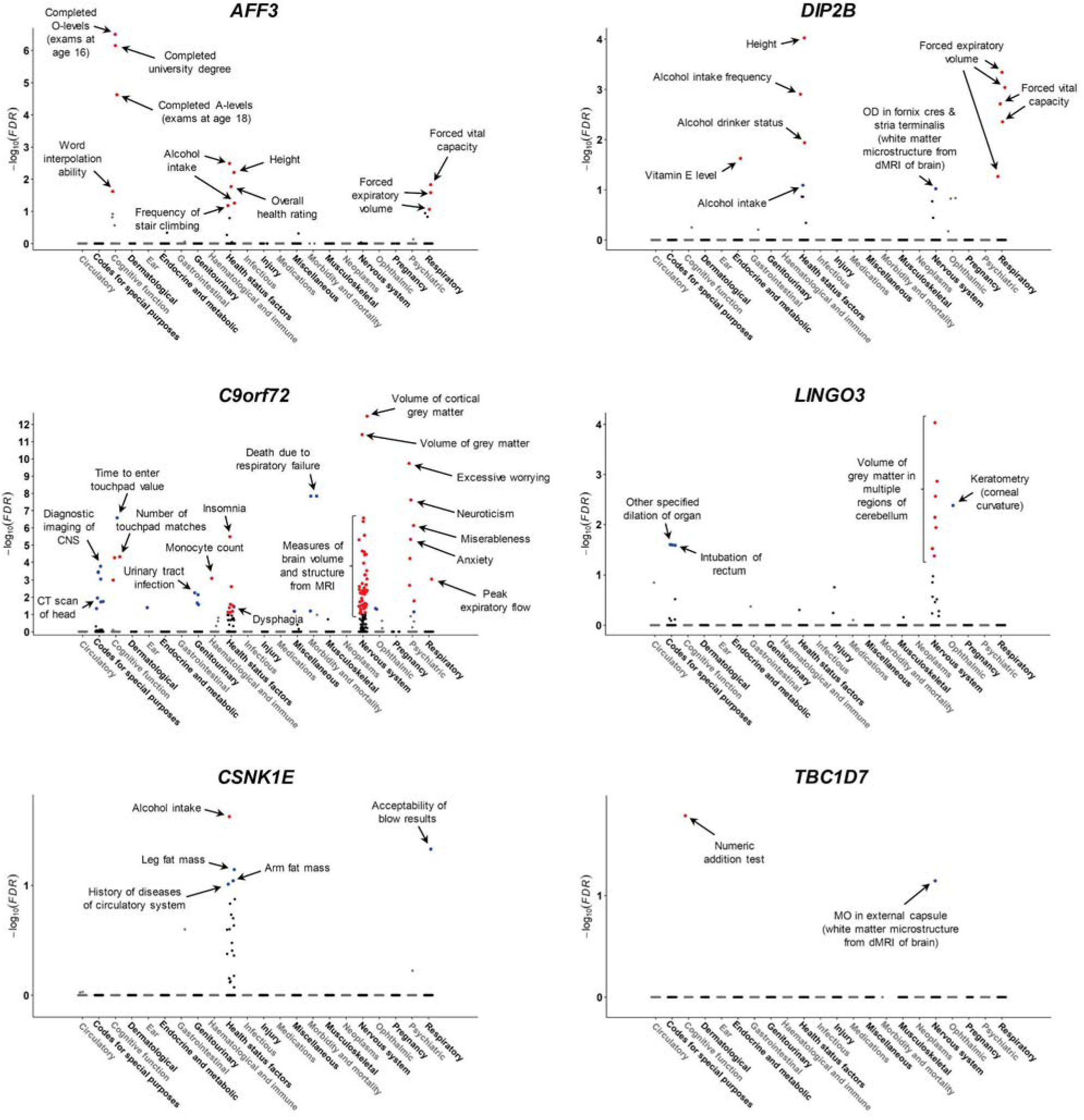
Results of phenome-wide association analysis for expansions of six GC-rich tandem repeats. Traits are grouped into physiological categories (x-axis) and we plot the -log_10_ FDR-corrected q-value (y-axis). Significant associations (FDR q<0.1) are individually labeled and shown in color to indicate directionality of effect (blue for positive associations, red for negative associations). Note that in some cases the trait names shown have been modified for clarity/brevity. Where a TRE:trait pair was significant under multiple definitions for identifying TREs (see Methods), we show only the most significant result for that TRE:trait pair. Results for additional TREs are shown in Supplementary Figure 7, with complete data in Supplementary Tables 6 and 7. Abbreviations: dMRI, Diffusion Magnetic Resonance Imaging; CT, Computed Tomography; CNS, Central Nervous System; MO, Mode from dMRI (a measure of microstructure of white matter).

In addition to these known pathologies, we also identified many other significant associations with TREs. For example, a GCC expansion in an alternative promoter of *AFF3*, which was previously suggested as potentially linked with neurodevelopmental problems^24^, showed a strong association with reduced educational attainment, with a 2.1-fold lower probability of obtaining O-levels (exams normally taken at age 16) (p=9.4x10^- 11^) and a 2.4-fold lower probability of obtaining a university degree (p=9.6x10^-11^). Individuals with this *AFF3* TRE also showed significantly reduced height, lung function, physical activity and reduced alcohol use as adults, had lower fluid intelligence and word interpolation abilities and self-reported worse overall health compared to controls, indicating negative effects of this expansion on multiple physical and mental traits.

Similarly, a GGC TRE in the 5’UTR of *DIP2B* (estimated frequency ∼1 per 1,000), which is known to cause silencing of *DIP2B* and the FRA12A folate-sensitive fragile site and was previously identified in several individuals with neurocognitive problems^25^, was associated with significantly reduced height, lung capacity, vitamin E levels, altered patterns of alcohol use and changes in the microstructure of brain white matter from diffusion MRI, accompanied by a nominal association (p=0.0002) for lower age of completing full time education.

Other notable signals we identified (Figure 3, Supplementary Figure 7) include:

i. Expansions of a CGG repeat in the 5’UTR of *CBL* (estimated frequency ∼1 per 6,000) associated with increased blood platelet levels (p=1.1x10^-14^). Consistent with the known link between coding variants in *CBL* and juvenile myelomonocytic leukemia^39^, we also observed that carriers of this TRE showed a nominal association with chemotherapy treatment for neoplasms (p=0.0005).
ii. Expansions of a GCC repeat in the 5’UTR of *LINGO3* (estimated frequency ∼1 per 850) associated with reduced volume of gray matter in multiple regions of the cerebellum (peak p=8.8x10^-9^), suggesting potential links of this TRE with neurodegeneration.
iii. Expansions of a GCC repeat in the 5’UTR of *ADARB1* (estimated frequency ∼1 per 10,000) associated with recent changes in speed/amount of moving or speaking (p=3.3x10^-7^), suggesting potential links with depression or late-onset neurological disease.
iv. Expansions of a degenerate GCY repeat in the 5’UTR of *TBC1D7* (estimated frequency ∼1 per 4,000) associated with significantly reduced performance on the numeric addition test (p=9.2x10^-6^) and altered microstructure of brain white matter from diffusion MRI (p=2.1x10^-5^).
v. Expansions of a CGC repeat in the 5’UTR of *CSNK1E* (estimated frequency ∼1 per 4,000) associated with reduced alcohol intake, increased body fat and increased incidence of diseases of the circulatory system.
vi. Expansions of a CGC repeat in the 5’UTR of *BCL2L11* (estimated frequency ∼1 per 20,000) associated with blood eosinophil and neutrophil levels.

Underlying data distributions for significant associations are shown in Supplementary Figures 8-16.

### Replication of AFF3 expansions with reduced educational attainment in the All of Us cohort

We genotyped the GCC repeat in *AFF3* in 85,512 unrelated individuals of European, African and Latino/Native American ancestries sampled from the population of the USA and utilized data for highest education level attained. Consistent with our findings in the UKB, we observed a significantly reduced probability for *AFF3* expansion carriers to attain a college degree (peak p=0.0006, odds ratio 0.53) (Supplementary Table 8).

### AFF3 expansions are enriched in unsolved probands with intellectual disability in The 100,000 Genomes Project

The 100,000 Genomes Project (100kGP)^40^ recruited 90,190 individuals with a range of rare diseases of unknown etiology or cancer, including 7,871 probands with a variety of neurodevelopmental disorders (https://re-docs.genomicsengland.co.uk/current_release/). Illumina GS was performed on each individual and, as of January 31^st^ 2023, pathogenic or likely pathogenic mutations had been identified in 19% of those recruited. Given the observed association in UKB between a TRE in *AFF3* and educational attainment, we hypothesized that TREs of *AFF3* might represent the pathogenic mutation in some patients with neurodevelopmental disorders. We genotyped the GCC TR in *AFF3* in this cohort and compared the frequency of long alleles in the 6,371 unsolved cases with a primary referral reason of intellectual disability to 8,794 controls, approximately matched for ancestry (Supplementary Figure 17, Supplementary Table 9). While the distributions of *AFF3* repeat alleles in the common polymorphic range (99% of control alleles were ≤38 copies) were indistinguishable between cases and controls, we observed a significant excess of long *AFF3* alleles in probands in whom no other pathogenic mutation had been identified (p=0.002, 3.9-fold enrichment for alleles ≥61 copies) (Figure 4), indicating this TRE as the likely pathogenic mutation in these individuals. As a further indicator of causality, the enrichment of *AFF3* expansions in idiopathic neurodevelopmental disorders occurred exclusively in probands where no other presumptively pathogenic mutation had been identified. Considering expanded *AFF3* alleles as those genotyped by ExpansionHunter as ≥60 copies of GGC (Figure 1), 17 of 6,371 unsolved probands had *AFF3* expansions, compared to none of the 1,500 probands in whom other pathogenic mutations had been identified by GS (p=0.056, Fisher’s exact test). Thus, our observations indicate that GCC expansions of *AFF3* represent the causal genetic lesion in ∼0.3% of patients with neurodevelopmental disorders, although this is potentially an underestimate due to the limitations of accurately sizing long GC-rich TR alleles from Illumina GS data^20,21^.

**Figure 4.**
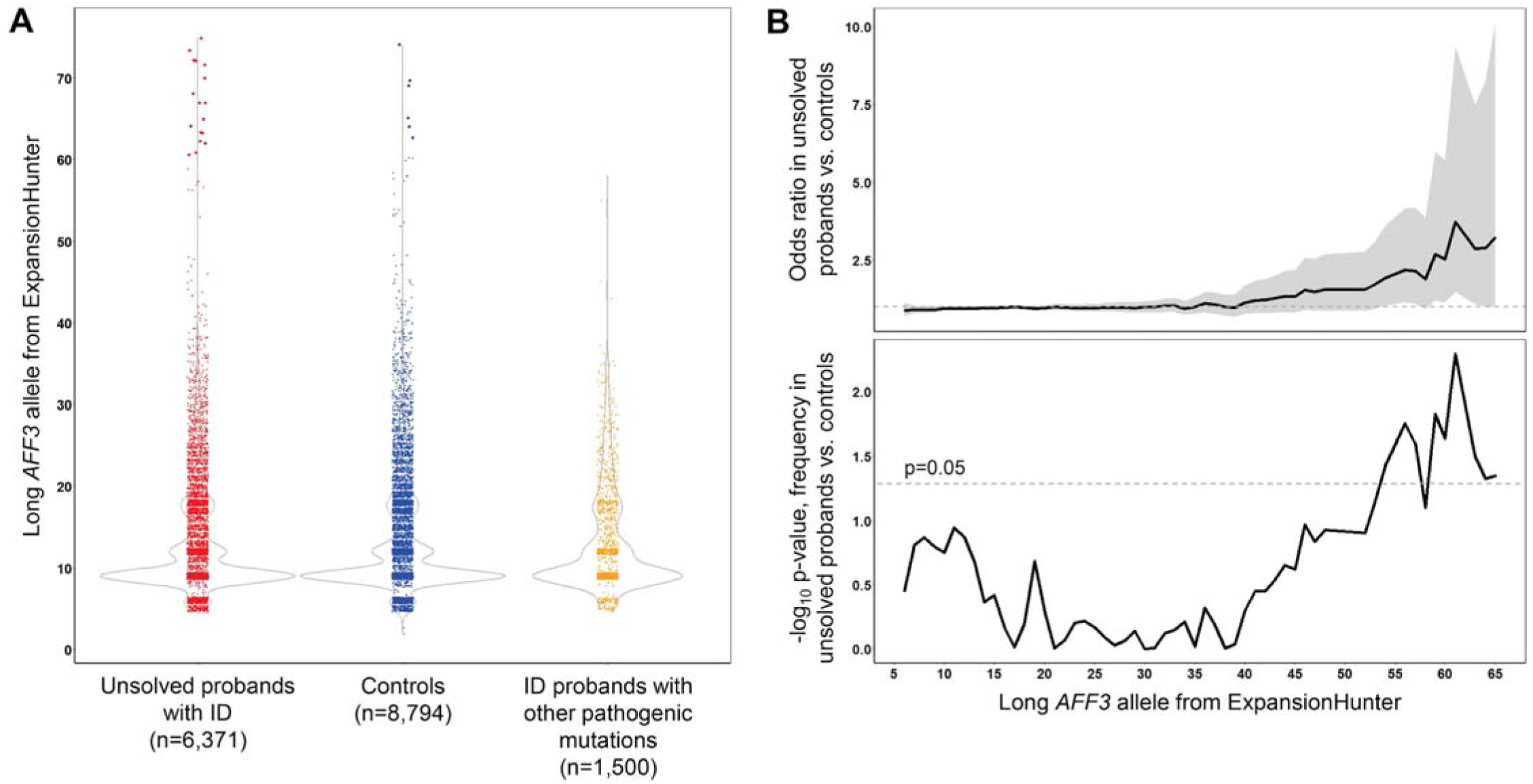
*AFF3* expansions are enriched specifically in unsolved probands with intellectual disability in The 100,000 Genomes Project. (A) Distributions of *AFF3* GCC TR alleles observed in previously unsolved probands with ID (red), controls (blue) and ID probands with other pathogenic or likely pathogenic mutations considered causal for their phenotype (orange) within the 100kGP cohort. TR alleles >60 copies are plotted as larger points. **(B)** Odds ratio and associated p-value for the presence of long *AFF3* TR alleles in 6,371 unsolved probands with a primary referral reason of intellectual disability compared to 8,794 controls. Peak enrichment occurs for alleles of ≥61 copies (p=0.002, 3.9-fold enrichment). In the upper panel, the gray shaded region represents the 95% confidence interval of the odds ratio, while the horizontal dashed line represents no enrichment (odds ratio=1). In the lower panel, the horizontal dashed line represents the significance threshold of p=0.05. To maintain statistical robustness, for the purposes of this plot, alleles ≥65 copies were combined as a single group.

Based on data provided by the referring clinical centers, in addition to the primary referral reason of intellectual disability/global developmental delay, probands with *AFF3* expansions presented with a variety of other recurrent phenotypes. Of these, speech and motor delays were observed in 59%, seizures in 41%, behavioral disturbances in 35%, generalized hypotonia in 24% and a variety of dysmorphic features and congenital anomalies at lower frequencies (Supplementary Table 10).

### Long read sequencing of trios with AFF3 expansions

We performed Pacific Biosciences HiFi GS in two trios with putative expansions of the GCC repeat in *AFF3* where the proband presented with intellectual disability. In both trios, we observed heterozygous expanded *AFF3* alleles composed of apparently pure GCC motifs in both the proband and one parent (Figure 5). All expanded alleles showed evidence of somatic mosaicism for length (Supplementary Figure 18) and varying levels of DNA methylation, ranging from 35% in an unaffected mother who carried a consensus 133-copy allele to 94% in their affected son who carried an 822-copy allele. Thus, in this trio, the mother’s 133 copy allele underwent *de novo* expansion to 822 copies upon transmission to the proband accompanied by increased methylation. In another trio, the unaffected father carried a highly (94%) methylated 424 copy allele that underwent contraction to 157 copies upon transmission to the affected proband, with slightly lower DNA methylation levels (75%). All four of these *AFF3* alleles were considerably longer than observed in 1,027 population controls with HiFi GS from the All of Us cohort and, as the father in trio 1 had no history of reduced cognitive function, strongly indicates variable penetrance associated with *AFF3* expansions.

**Figure 5.**
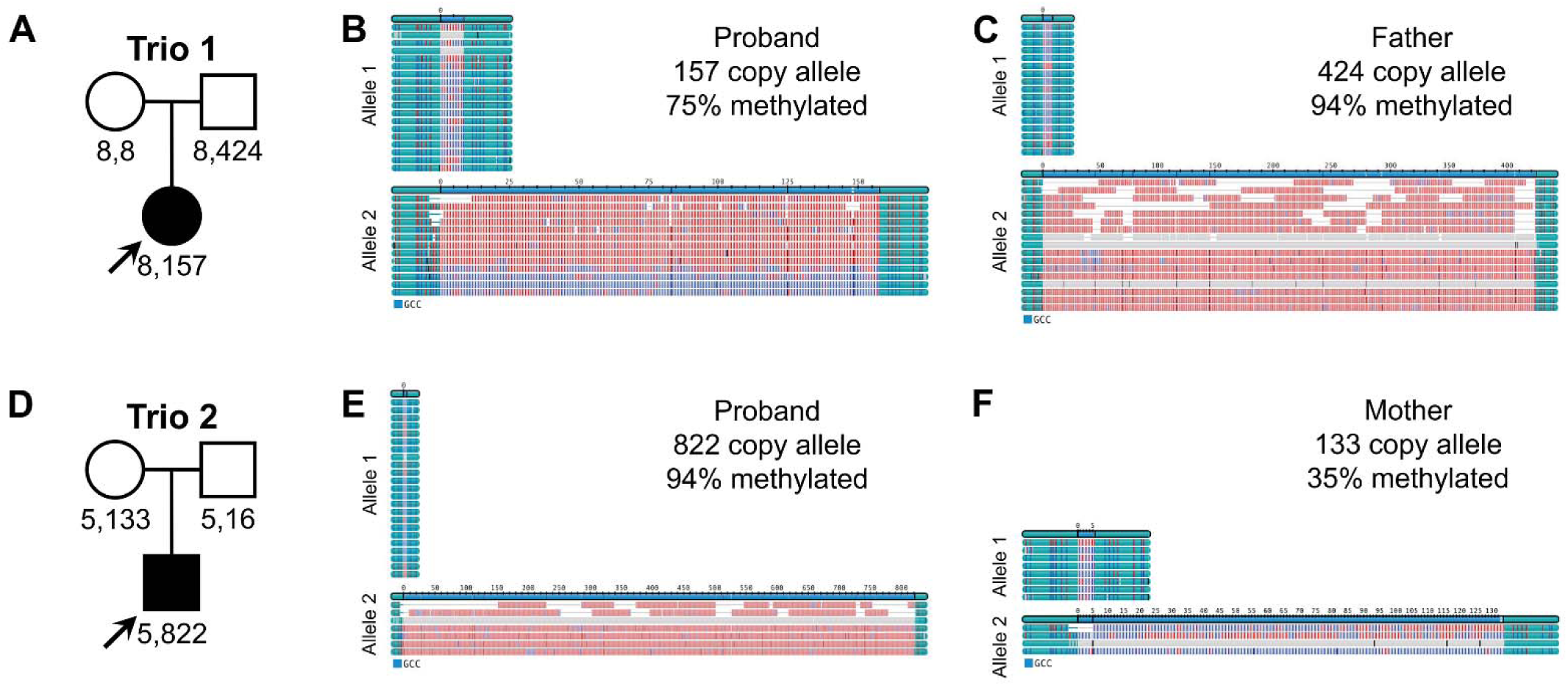
Pacific Biosciences HiFi GS data of two trios carrying *AFF3* expansions. **(A and D)** Pedigree structure of each trio showing *AFF3* alleles observed in each family member. **(B, C, E and F)** Consensus size and methylation status of expanded *AFF3* alleles in the proband and transmitting parent, as reported by TRGT. Methylated GCC motifs are shown in red, unmethylated in blue, flanking sequences in cyan.

Using the genome-wide data, we performed transmission analysis of 937,122 TRs in each trio using TRGT-denovo, a tool designed to detect *de novo* TR mutations in trios. In both trios, the GCC expansion in *AFF3* was the top ranked *de novo* repeat allele in the proband’s genome based on change in size versus the parental alleles, providing further evidence to support their pathogenicity (Supplementary Table 11).

### Relative impact and modifiers of AFF3 expansions on educational attainment

To assess the relative effect size of the *AFF3* TRE on educational attainment in comparison to other mutations that impact neurodevelopment, we used read depth from GS to identify several recurrent microdeletion/duplications in the set of 168,641 UKB individuals used for PheWAS (Supplementary Figure 19). Among these, expansions of *AFF3* exert a negative effect on educational attainment that has an effect size comparable to that seen in carriers of del(22q11.21), del(1q21.1) and del(15q13.3) (Figure 6).

**Figure 6.**
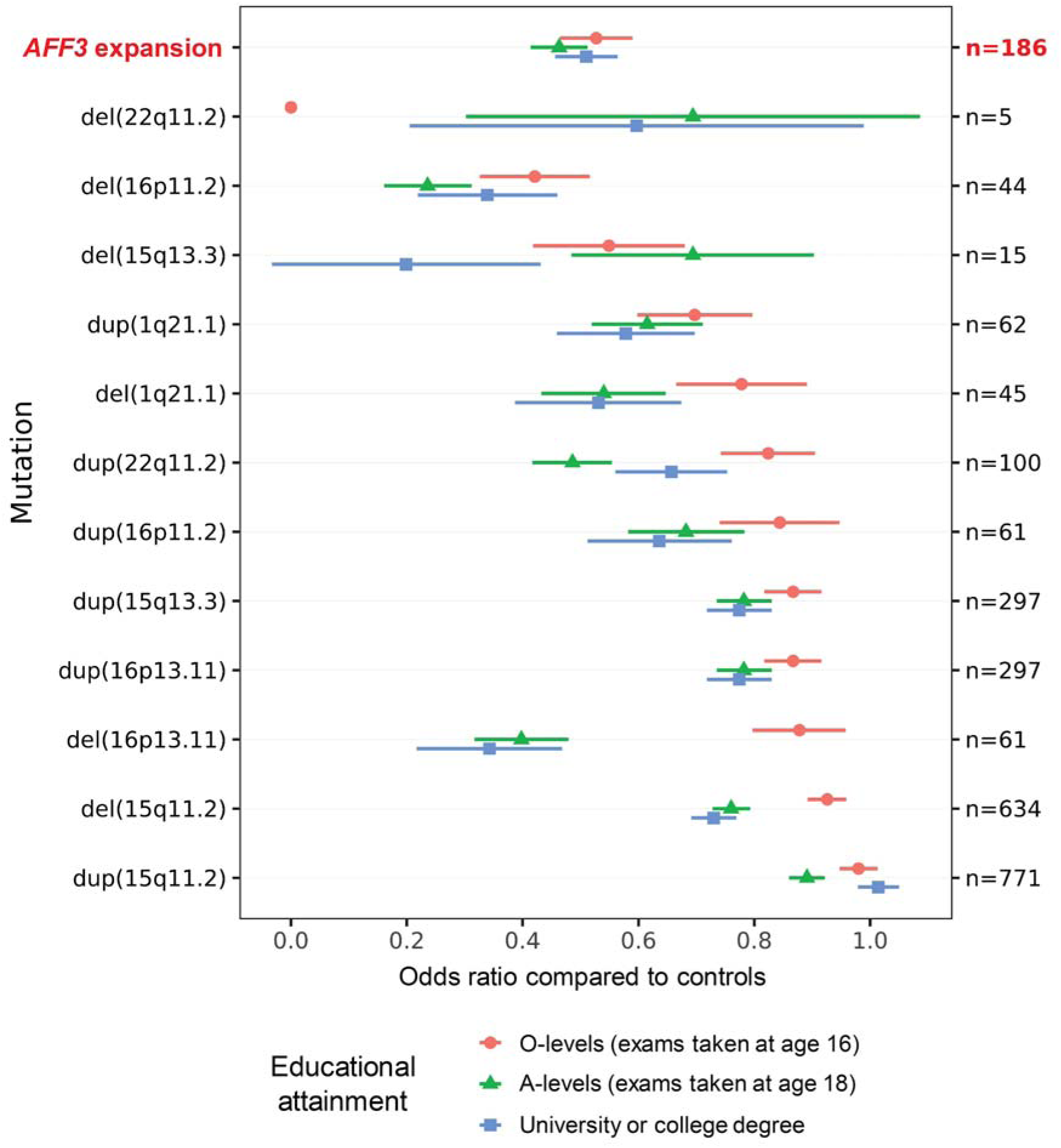
Relative effects of the *AFF3* expansion on educational attainment compared to recurrent microdeletions/duplications. Points show the odds ratio of each group, with horizontal lines indicating 95% confidence intervals. CNVs are ordered by odds ratio for completing O-levels (exams taken at age 16) compared to controls that do not carry any of these mutations. The number of individuals with each mutation type among the 168,641 UKB samples assayed is shown on the right vertical axis. The number of methylated *AFF3* expansions is based on an estimated population prevalence of 1 per 862, with effect sizes on educational attainment based on the 186 individuals with the largest *AFF3* alleles in the UKB individuals utilized for PheWAS.

Given the apparent phenotypic heterogeneity among individuals with *AFF3* expansions, with some obtaining university degrees, we investigated possible modifiers of penetrance. Using a large set of SNVs identified from GWAS^41^, we calculated a PRS for educational attainment in UKB participants. Among individuals with *AFF3* expansions, we observed significantly higher PRS in those who obtained A-levels (exams normally taken at age 18) compared to those that did not (p=0.02) (Supplementary Figure 20). Partitioning individuals with *AFF3* expansions based on quintiles of PRS showed a clear influence of genetic background on educational attainment, with individuals in the upper quintile of PRS showing an ∼2-fold higher likelihood of obtaining A-levels compared to those in the bottom quintile (Supplementary Figure 21). Similar results were also observed for several recurrent microdeletions/duplications. These observations strongly suggest that the negative effects of *AFF3* expansions and other recurrent CNVs on educational attainment are modified by polygenic background.

### Population prevalence of methylated AFF3 expansions

As it can be difficult to accurately estimate the true size of expanded GC-rich TRs from Illumina GS, in order to provide a more accurate estimate of the frequency of methylated *AFF3* expansions that cause the FRA2A fragile site, we utilized DNA methylation data from 32,776 individuals profiled using the Illumina 450k and 850k arrays^19,42^, identifying 38 individuals with hypermethylation of the *AFF3* locus (Supplementary Figure 22). Thus, presuming these hypermethylation events correspond to the presence of an underlying TRE, we estimate the population prevalence of methylated *AFF3* expansions as ∼1 per 862 individuals (95% confidence interval, 1 per 654 to 1,266).

### SNVs that associate with presence of the AFF3 repeat expansion match those found by GWAS

Multiple prior GWAS have reported SNVs at the *AFF3* locus that show significant associations with intelligence, cognitive ability and educational attainment^43^. Based on the observation that, at some TRE loci, local SNVs show significant associations with the presence of a TRE^44–50^, we hypothesized that the GCC TRE in *AFF3* might be the causal variant that underlies these GWAS signals. Consistent with this, we identified hundreds of SNVs located within the *AFF3* locus that show strong association with presence of an expanded GCC TR allele and observed that many of these are the same SNVs that have been previously identified by GWAS for intelligence, cognitive ability and educational attainment (Figure 7, Supplementary Table 12). We repeated analysis of the *AFF3* TRE with educational attainment after conditioning on the lead tag SNV (rs60473748) and still observed a highly significant association (peak p=7.9x10^-11^ for obtaining a university degree), indicating that the effect of the *AFF3* TRE is independent of local SNVs. These observations provide further evidence for the role of the *AFF3* TRE in neurodevelopment and indicate that a subset of GWAS signals are likely driven by underlying TR variants that preferentially occur on specific founder haplotypes.

**Figure 7.**
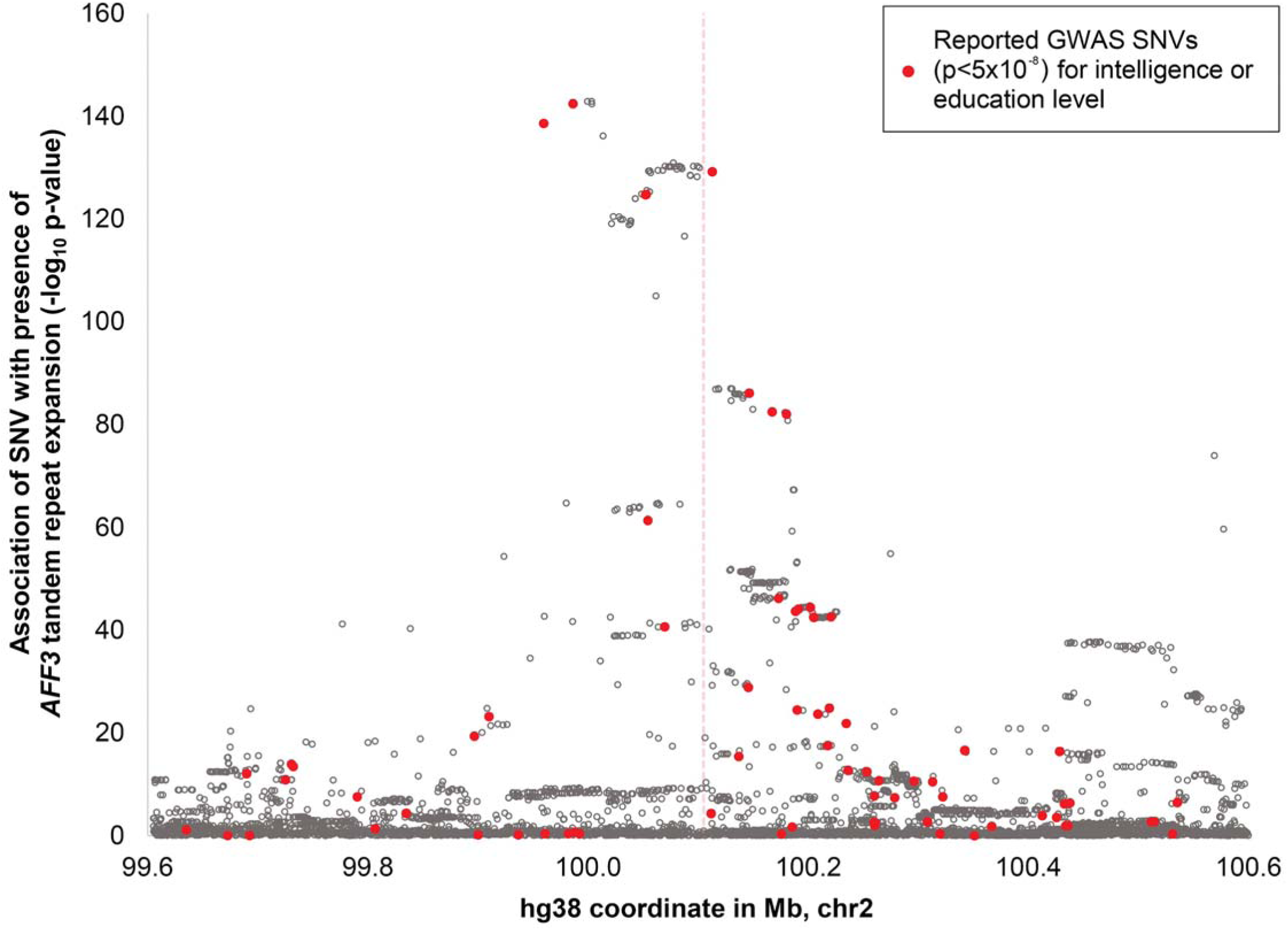
SNVs that associate with presence of the *AFF3* repeat expansion are the same as those identified in prior GWAS of intelligence, cognitive ability and educational attainment. We identified SNVs that associate with the *AFF3* TRE in the UKB cohort and joined these with SNVs for intelligence, cognitive ability or education level reported in the GWAS catalog (shown as red points). The vertical dashed line in the center of the plot indicates the position of the GCC TR within *AFF3*.

## DISCUSSION

In 1991, the recognition of a methylated repeat expansion in *FMR1* as the causal mutation underlying fragile X syndrome represented a new paradigm for the genetic basis of human disease^2,51^. However, the inherent technical difficulties of genotyping long TR alleles has meant that progress in elucidating the wider role of TREs in human traits over the past three decades has been relatively modest^4^. Here, we performed a three-part study in which we first used a combination of epigenome profiling and TR genotyping to identify 24 methylated TREs in the human genome which resemble the TRE that causes fragile X syndrome, then performed population-scale PheWAS to identify the effects of these TREs on diverse human traits and finally tested their contribution to rare disease in a cohort of patients who remained refractory to genetic diagnosis despite undergoing GS. In addition to several that were previously described, we identified multiple methylated TREs that had not been previously recognized and found that many of these exert diverse effects on human traits.

Of note, analysis in the UKB showed that expansions of a GCC motif within *AFF3* have a strong negative effect on educational attainment, cognitive function and multiple measures of general health. Comparison with other recurrent CNVs showed that the effect of these *AFF3* expansions on educational attainment are similar in magnitude to those caused by several clinically-significant microdeletions that are a frequent cause of neurodevelopmental anomalies, such as del(22q11.2)^52^, del(1q21.1)^53,54^ and del(15q13.3)^55,56^. As evidenced by their presence in both unaffected carrier parents and dozens of individuals in the UKB and All of Us cohorts who completed university education, the *AFF3* TRE shows incomplete penetrance and we showed that this phenomenon is modified by polygenic background. It should be noted that the UKB recruited individuals from an apparently healthy population, creating an ascertainment bias that will lead to underestimates in the prevalence of more severe neurocognitive deficits and morbidities that may be associated with a mutation. As such, it is likely that our results from the UKB underestimate the true impact of this TRE on cognitive function. In contrast, patients recruited by the 100kGP represent individuals that had been referred to clinical genetics centers but in whom prior genetic testing had failed to identify any causal mutations and thus represent a sample of individuals that tend to exhibit more severe developmental anomalies. Despite this, results from these two populations converged, showing that these *AFF3* TREs represent a significant cause of developmental delay. We estimate the prevalence of methylated *AFF3* expansions as ∼1 per 850 individuals in the general population, a frequency that is 5- to 10-fold higher than the estimated prevalence of CGG expansions within *FMR1* that cause fragile X syndrome^1,57^ and 3- to 8-fold higher than recurrent microdeletions/duplications of 1q21.1, 15q13.3 and 16p11.2^58^. We, therefore, propose that the *AFF3* TRE likely represents a relatively common cause of developmental anomalies and recommend that testing for this TRE should be considered in individuals with unexplained developmental delay.

ExpansionHunter genotypes for GC-rich TREs exceeding the read length of Illumina GS are estimates that often show considerable inaccuracy^20,21^, which creates several limitations: (i) We observed that size estimates produced by ExpansionHunter for GC-rich TREs are consistently shorter than the true allele sizes from long-read GS. (ii) As a result, using genotypes from Illumina GS alone, it is difficult to distinguish large expansions from shorter premutation alleles, or to accurately estimate the prevalence of the TREs we studied. (ii) As such, our analysis potentially underestimates the true prevalence of *AFF3* expansions as a cause of intellectual disability and it is possible that some of the associations we identified may be with unmethylated premutation alleles. (iii) Where an expanded *AFF3* allele was also detected in one parent of a proband, using short read GS alone, we were unable to reliably distinguish whether these were premutation alleles that underwent further expansion into the pathogenic range upon transmission, or discern whether there was any parental bias in transmission to affected offspring^59^. Studies of larger numbers of pedigrees using more accurate genotyping methods will be needed to examine these issues. Furthermore, it should be noted that at some loci, genotypes from ExpansionHunter can include a high frequency of false-positive TREs and thus require quality control to ensure accurate identification of TREs.

The original report that identified a recurrent TRE at *AFF3* underlying the FRA2A fragile site showed that these expansions also caused methylation and allelic silencing of *AFF3*^24^. Our data support the notion that methylated and transcriptionally silent full mutations of *AFF3*, rather than shorter unmethylated premutation alleles, are causal for intellectual disability. However, while this potentially implicates haploinsufficiency of *AFF3* as the underlying pathogenic mechanism, current evidence is unclear. Missense mutations of *AFF3* have been identified in KINSSHIP syndrome^60^, which includes a spectrum of anomalies not observed in carriers of the *AFF3* GCC expansion. However, variants causing KINSSHIP syndrome all occur within a single protein domain thought to be important for the interaction of AFF3 with its binding partners, suggesting the syndrome results from a specific dysregulation of the AFF3 pathway. As the GCC repeat expansion in *AFF3* is intronic, an alternative possibility is that expanded alleles may exert pathology through gain-of-function mechanisms such as RNA toxicity or RAN-translation^61,62^. Further work will be needed to better elucidate the underlying pathological mechanisms of this TRE.

Although we utilized data from >168,000 deeply phenotyped individuals in our PheWAS, statistical power to detect associations was relatively limited for many of the loci and phenotypes we tested as most TREs and many of the traits we studied are rare. Despite this, it is notable that a high proportion of the TREs tested here exhibited associations with human traits. In an era where SNV-based GWAS are one of the most common methods utilized for investigating the genetic basis of human disease, our study highlights the role of more complex recurrent variants that are poorly assayed by GWAS as potential contributors to the “missing heritability” of the human genome^63,64^. Emphasizing this, by directly studying TRs, we were able to identify the TRE at *AFF3* as showing a much stronger effect on educational attainment than nearby SNVs reported in prior GWAS. We postulate that other loci identified by GWAS may also be attributable to underlying TR variants, a possibility that should be considered in fine mapping studies. We anticipate that the increasing availability of large Biobanks with both GS and extensive phenotype data will enable many novel insights into the role of TREs in human disease and variation.

## ONLINE METHODS

### Quality control and data processing of methylation data

For the identification of epivariations, we utilized methylation data from 8,371 individuals from the Project MinE cohort^42^. Informed consent for genetic research was obtained from all participants, approved by the Trent Research Ethics Committee 08/H0405/60. DNA methylation profiles were generated using either the Illumina HumanMethylation450 BeadChip (henceforth called “450k array”) (n=4,474) or the Illumina Infinium MethylationEPIC BeadChip (henceforth called “850k array”) (n=3,897), using DNA extracted from peripheral whole blood. We removed any sample with >1% of autosomal probes with detection p-value >0.01 and performed Principal Component Analysis (PCA) based on β-values of all probes located on chr1, removing outlier samples. Data were then normalized, as described previously^65,66^. Briefly, raw signal intensities were subjected to color correction, background correction and quantile normalization using the Lumi package in R^67^ and the normalized intensities converted into β-values, which range between 0-1, representing the methylation ratio at each measured CpG. In order to correct for inherent differences in the distribution of β-values reported by Infinium I and Infinium II probes, we applied BMIQ^68^. Data from 450k and 850k arrays were normalized independently. After normalization, we estimated the major cellular fractions comprising each blood sample directly from β-values using the method described by Houseman^69^ and removed outlier samples, defined as those that showed cellular fractions either ≥99^th^ percentile +2%, or ≤1^st^ percentile -2% of any cell type. After all quality control and filtering steps, 7,447 samples were processed to identify epivariations, comprising 5,403 individuals with a diagnosis of amyotrophic lateral sclerosis (ALS) and 2,044 controls. Of these, 5,750 also had available GS data.

### Identification of rare epigenetic variants

In order to identify rare epigenetic variants, also termed differentially methylated regions (DMRs), we utilized a sliding window approach to compare individual methylation profiles of a single sample against all other samples. This process was performed separately using data from the 450k and 850k array platforms. We defined DMRs as regions of outlier methylation represented by multiple independent probes using the following parameters:

Hypermethylated DMRs: Any 1 kb region with at least three or more probes with β-values ≥99.5^th^ percentile plus 0.15 and contains at least three consecutive probes with β-values ≥99.5^th^ percentile. In addition, we required that the minimum distance spanned by probes that were ≥99.5^th^ percentile was ≥100 bp.

Hypomethylated DMRs: Any 1 kb region with at least three or more probes with β-values ≤0.5^th^ percentile minus 0.15 and contains at least three consecutive probes with β-values ≤0.5^th^ percentile. In addition, we required that the minimum distance spanned by probes that were ≤0.5^th^ percentile was ≥100 bp.

As the presence of an underlying homozygous deletion at a probe binding site can result in spurious β-values^65^, we removed any DMR call in which the carrier individual reported one or more probes within the DMR with failed detection p-value (p>0.01). Finally, we removed samples with an unusually high number (n>20) of autosomal DMRs.

For the GCC repeat within *AFF3*, as only two probes on the 450k and 850k arrays show a clear increase of methylation levels in individuals with an expanded repeat allele, this locus did not meet our stringent genome-wide discovery criteria for identifying DMRs. Therefore, for this locus, we considered hypermethylation events as those where the normalized β values of probes cg14267725 and cg23118464 were both >99.8^th^ percentile of their respective population distributions.

For the GGGGCC repeat within *C9orf72*, as the cohort is comprised of 72% of individuals with ALS and, thus, is highly enriched for carriers of expanded *C9orf72* TR alleles, we considered hypermethylation events at this locus as those where the normalized β values of at least two probes within 1kb of the TR were each >99^th^ percentile of their respective population distributions.

### Identification of tandem repeat expansions in the Project MinE cohort

In order to identify putative TREs potentially linked with rare hypermethylation events, we performed two analyses. First, we utilized ExpansionHunter DeNovo^70^ to identify genomic loci containing TR alleles ≥150 bp in length. The regions output from ExpansionHunter DeNovo were overlapped with DMRs identified by methylation profiling (extended by ±500 bp) and, where these regions intersected within the same sample, they were taken forward for targeted genotyping using ExpansionHunter (v3.2).

In addition, we utilized a set of highly polymorphic TRs that showed ≥10 different alleles in the population based on profiling with HipSTR^71^. Based on the observation that a high rate of polymorphism in the population is a very strong predictor of unstable TRs that also undergo occasional extreme expansions^19,72^, we reasoned these would be strongly enriched for possible TREs. These putatively unstable TRs were intersected with hypermethylated DMRs identified by methylation profiling (extended by ±500bp) and any overlapping TRs were then genotyped using ExpansionHunter. To identify TREs that associate with local DNA methylation, at each locus we performed 100 million permutations to assess the probability of observed distribution of methylated TR alleles. In each permutation, we randomly assigned hypermethylated samples and compared the mean TR length of individuals with hypermethylation with the observed mean, generating an estimated p-value based on the fraction of simulations where the permuted mean exceeded the observed value.

### Long read sequencing of putative tandem repeat expansions

Pacific Biosciences long insert libraries with the addition of barcodes were prepared for samples with epivariations at *DPYSL4* and *FAM193B*, the two samples mixed at equimolar amounts and sequenced on a single 8M SMRT cell with the Pacific Biosciences Sequel II system in continuous long read (CLR) mode. Subreads were aligned to the hg38 human reference genome using pbmm2 v1.0.0 with default parameters.

We sequenced a sample with hypermethylation of the *ELF1* promoter using Oxford Nanopore Technology (ONT) to a mean depth of 12x coverage. Reads were mapped to the hg38 human reference genome using minimap2 (v2.7)^73^ and the bam file sorted and indexed using samtools (v1.7)^74^. To estimate methylation levels on normal and expanded CGG repeat alleles separately, we first phased the reads using SNVs within the reads. Using nanopolish (v0.10.2)^75^, we created index files to link reads with their signal level data in FAST5 files, followed by estimation of DNA methylation status at each CpG located within 2 kb of CGG TRs, requiring a minimum log likelihood ratio ≥2.5 at each site.

### Expression studies of genes with methylated repeat expansions in the GTEx cohort

We utilized published DNA methylation data generated using the Illumina 850k array from 9 tissues and 424 individuals in the The Genotype-Tissue Expression (GTEx) cohort^34^ to first identify outlier hypermethylation events at the 24 methylated TREs that were identified in Project MinE samples. We then utilized corresponding RNAseq data from 948 individuals published on the GTEX portal (Version 9, https://www.gtexportal.org/) to compare normalized expression levels (TPM) and WASP-corrected allele-specific expression (ASE) data of the relevant gene in carriers of each TRE to other tissue-matched samples in the GTEx cohort. We performed permutation analysis to estimate the probability of seeing the observed distribution of expression ranks or allelic bias in carriers of hypermethylation compared to that in the overall cohort. Here, we performed 10,000 random permutations of the observed expression ranks or allelic ratios in each hypermethylation carrier for all available tissues against the rest of the GTEx cohort.

### Identification of tandem repeat expansions in the UK Biobank

Collection of the UKB data was approved by the Research Ethics Committee of the UKB obtained under application 32568. All study participants provided informed consent and the protocols for UKB are overseen by The UKB Ethics Advisory Committee, see https://www.ukbiobank.ac.uk/ethics/. From the set of 200,000 individuals with Illumina 150 bp paired-end whole GS data, we defined 188,915 individuals of European ancestry using an approach based on principal component analysis of a set of ∼64,000 high quality linkage disequilibrium (LD) pruned SNVs with minor allele frequency (MAF) >5%, as described previously^76^. In brief, GCTA (v1.93) was used to calculate the first 40 Principal Components (PCs), which were used to predict the ancestry of each individual. Since only a small minority of individuals recruited to the UKB cohort are of non-European ancestry and are, therefore, highly under-powered in PheWAS, we focused our analysis on those individuals of European origin. We first selected all samples predicted as European ancestry and recalculated PCs without use of a reference population using one third of individuals, before projecting the remaining two thirds of individuals onto this. We removed outlier samples using within group PCA. In addition, we kept only those samples with self-reported ancestry as “White”, “British”,”Irish”,”Not listed in table”, “Any other white background”, “Other ethnic group”, or “Prefer not to answer”. Further sample-level filters were applied as follows:

1. Using pairwise kinship coefficients provided by the UKB, for samples with 2^nd^ degree relationships or higher (kinship coefficient >0.0883) we retained only a single unrelated individual.
2. We removed 199 individuals with predicted sex chromosome aneuploidy.
3. We removed 241 individuals who we were notified had withdrawn their consent for inclusion in the UKB study.

After these quality control (QC) steps, we retained data for 168,641 unrelated individuals of European ancestry. Using the UKB DNAnexus Research Analysis Platform, we performed genotyping of 28 GC-rich TRs from the GS data with ExpansionHunter (v5)^77^ (Supplementary Table 13). From the resulting diploid genotypes output by ExpansionHunter, we utilized data only for the longer of the two alleles per sample at each locus, discarding data from the shorter allele. To avoid artifacts due to possible technical effects on genotypes, samples were then separated by sequencing center (deCODE and Sanger Center) and, for loci on the X chromosome, samples were further separated by sex. Based on the set of alleles observed at each TR locus per sample group, we calculated population statistics and used these for the determination of putative expanded alleles using six levels of stringency (named Very low, Low, Medium, High, Very High and Extremely High), each of which utilized the absolute and percentile allele size, difference from the population median and outlier probability based on the z-score (Supplementary Table 4). An example of the output of these different stringencies of outlier calling are shown in Supplementary Figure 5.

To reduce false positive calls for TREs that often result from low read depth or poor-quality alignments of Illumina reads over TR regions^78^, we developed a Random Forest classifier to recognize low-confidence TRE calls based on characteristics of the TR locus and the output metrics supporting each genotype generated by ExpansionHunter. We first trained this tool on a set of ∼3,500 putative TREs from ExpansionHunter for which we performed manual curation of read pileup plots to determine true and false positives. For each putative TRE, we generated a visualization of reads overlapping the TR locus using GraphAlignmentViewer (https://github.com/Illumina/GraphAlignmentViewer) and by manual inspection classified these as either true or false based on the number and quality of the read alignments supporting the putative TRE. In general, TREs considered as true were supported by multiple reads with few mismatches, while those considered as false were often supported by one or very few reads with multiple mismatches in the alignment (Supplementary Figure 23).

To assess the performance of the trained model, we analyzed data from 1,027 individuals from the All of Us cohort for which both Illumina short read GS and PacBio HiFi long-read GS are available. Here, for the 28 GC-rich TRs used in our PheWAS, we generated TR genotypes from both PacBio GS data using TRGT and from Illumina GS data using ExpansionHunter, with identical TR definitions used for both tools. This yielded a set of genotypes for the 28 TRs derived from both long and short-read GS in the same set of individuals. Presuming that the PacBio HiFi data represent a gold-standard “truth set” of TR genotypes, we used these to benchmark the performance of ExpansionHunter for identifying TREs, both with and without quality filtering using our Random Forest Classifier. We first filtered to retain only high-quality genotypes produced by each tool. We removed TRGT genotypes that were supported by only a single spanning read or which had purity scores <0.75. We then identified putative outlier TREs in both genotype sets based on our Low stringency criteria (Supplementary Table 4). Finally, for the ExpansionHunter genotypes, we applied our Random Forest classifier to identify TREs that were scored as false positives.

We then calculated several statistics from these two genotype sets at each of the 28 loci, summarized in Supplementary Table 14:

i. The Spearman rank correlation (R), which gives a summary of the overall similarity of the two sets of genotypes, but which is heavily weighted by the much larger number of common (non-expanded) TR alleles.
ii. To better describe how accurately rare TREs are identified by ExpansionHunter, we calculated the fraction of TREs called from the ExpansionHunter genotypes that were also called from the TRGT genotypes and vice versa. This was done from both raw genotypes and after quality filtering the TREs using our Random Forest classifier, thus allowing the definition of how often rare TREs identified by ExpansionHunter were validated with long-read data (the true positive rate), what fraction of TREs called by ExpansionHunter did not validate with long-read data (the false positive rate) and what fraction of TREs observed in the long-read data were not identified by ExpansionHunter (the false negative rate).

Overall, we observed excellent correlations between the two genotype sets across the 28 TR loci tested, with most values of R>0.9. Even for TRs where the R value was low, *e.g. DPYSL4* (R = 0.1), ExpansionHunter was still able to accurately identify 83% of TREs at this locus. Further, comparison of TREs called by ExpansionHunter at these 28 loci both before and after quality filtering with our automated classifier showed that this increased the true positive rate for TREs from 82% to 93% and reduced the false positive rate calls from 10% to 6%, although the false negative rate increased from 5% to 13%. Inspection of these false negatives produced by our Random Forest Classifier showed that nearly all were alleles where the span of the expanded TR tract was between 140bp and 160bp, *i.e.* approximately the same as the sequencing read length and which often have zero or just a single supporting in-repeat read and no mapped reads that completely span the entire repeat allele.

Based on this analysis, we applied the trained Random Forest classifier to screen all putative TREs identified in the UKB called using each of the six outlier criteria and any genotypes that were scored as false positive TREs were removed from further analysis. Following this automated quality filtering step, we defined each individual with a TRE that passed QC as “expanded” and defined all other individuals as “not expanded”.

### PheWAS analysis in the UK Biobank

In order to identify traits associated with the presence of TREs, we input the set of filtered binary genotype calls from the six stringencies used for identifying TREs (“expanded” versus “not expanded”) into association analysis. We utilized phenotype data for individuals in the UKB derived from a total of 10,615 ICD10 codes and quantitative, categorical and binary traits (Supplementary Table 15), accessed through UKB application number 82094. Before performing association testing, phenotype data were processed as follows:

1. For phenotypes with multiple categories, *e.g.* ICD10 codes, we considered each category as a separate binary trait. If a subject had multiple entries for the same category, they were assigned as “1” (*i.e.* positive for that trait) if any of the instances were positive. We removed from analysis any categories labeled as “Prefer not to answer”, “Unsure” or “Do not know”.
2. ICD9 and ICD10 codes were merged into one using General Equivalence Mapping (GEM) files downloaded from https://www.cms.gov/Medicare/Coding/ICD10. We restricted controls to subjects who did not have a positive diagnosis for that phenotype.
3. In addition to using each individual ICD code as a separate trait, ICD codes were also grouped into higher level codes to yield additional summary traits with increased sample size. For example, the ICD10 code for Schizophrenia is F20, which is composed of 10 different sub-codes (e.g. F20.0 “Paranoid schizophrenia:, F20.2 “Catatonic schizophrenia”, F20.9 “Schizophrenia, unspecified”, *etc*.), with each subcategory containing between 4 and 894 individuals. Here, we created a new summary code for “Schizophrenia_group” comprising all sub-categories under F20 which included 1,485 individuals who were positive for any subtype of schizophrenia. Similar groupings were performed for any higher level ICD code that comprised multiple subtypes.
4. For phenotypes with >2 categorical outcomes, each outcome was assigned an integer value and these were analyzed as quantitative traits. *e.g.* the phenotype “Alcohol intake frequency” included six categories: “Daily or almost daily”, “Three or four times a week”, “Once or twice a week”, “One to three times a month”, “Special occasions only” and “Never”. Here, each sample was assigned an integer value ranging from 1 to 6, with each value corresponding to the six ordered frequencies for this phenotype. Categorical traits with just two separate categories were considered as binary traits.
5. For quantitative traits with integer values, where a subject had multiple instances, we utilized the mean of all instances rounded to nearest integer.
6. For quantitative traits with continuous values, where a subject had multiple instances, we utilized the mean of all instances.

For the 1,316 quantitative traits analyzed, we applied a rank based inverse normal transformation and required a minimum sample size of 50 phenotyped individuals and standard deviation ≥0.001 to be included in association analysis. For the 9,299 binary traits analyzed, we required a minimum sample size of at least 25 individuals with the trait and a minimum of at least five individuals that carried the TRE to be included in the analysis.

Association analysis was performed using *REGENIE*^79^, incorporating covariates of sex, age, GS insert size and the top five principle components derived from analysis of SNVs to account for ancestry. For binary traits, we utilized the Saddle Point Approximation function to reduce the type I error rate. To avoid possible technical effects on TR genotypes, associations were performed separately in samples sequenced by deCODE and Sanger Center, before combining results using z-score based meta-analyses in *METAL*^80^. We required that associations showed the same direction of effect in both sub-cohorts. For loci on the X chromosome, samples were separated by both sequencing center and sex and associations performed separately in the four resulting sub-groups (sequencing center + sex) before combining results using *METAL*, requiring that at least three of the four sub-groups met the minimum sample size requirements as stated above and all showed the same direction of effect.

We applied multiple testing corrections using both a false discovery rate (FDR) and Bonferroni approach based on the total number of TRs and independent traits analyzed. However, given that many phenotypes in the UKB are highly correlated (*e.g.* neutrophil count and neutrophil percentage), it should be noted that these multiple testing corrections tend to be overly stringent. We considered associations significant at 10% FDR (FDR q<0.1).

For creating plots of grouped PheWAS results, we assigned each trait reported by the UKB into 22 different categories based on shared physiological systems, tests or treatments. To create these groupings, phenotype codes were mapped to ICD10 chapters as listed in Supplementary Table 1 of Wang et al.^81^. Phenotypes that did not have a category assigned with these annotations were further annotated using categories provided within the UKB Data Showcase. Finally, we applied manual curation, removing categories that were composed of a very small number of traits and making reassignments to improve the consistency of groupings.

### Replication analysis in All of Us

We performed genotyping of the *AFF3* TR in 106,708 individuals with Illumina GS data from the All of Us v7 data release using ExpansionHunter. Individuals with ancestry defined by All of Us as “East Asian”, “South Asian”, “Middle Eastern” or “Other” were removed due to low numbers. The remaining samples were divided into 12 different groups based on both ancestry (EUR, AFR, AMR) as determined by All of Us and sequencing center / date. While samples were sequenced at three different sequencing centers (Baylor, University of Washington and the Broad Institute), we observed a marked shift in GS insert size for samples sequenced at the Broad Institute prior to September 2019 compared to those sequenced September 2019 onwards and thus we sub-divided samples sequenced at the Broad Institute into two sub-groups based on this sequencing date. Identification of *AFF3* expansions was performed separately in each of the 12 groups using six different stringencies based on the long allele in each sample, as described above.

Before performing association analysis, we removed related individuals, defined by All of Us as those with kinship scores >0.1 and individuals with predicted sex chromosome aneuploidy based on genome-wide read depth analysis using mosdepth^76^. Of the remainder, 85,512 individuals had data for highest education level attained, comprising 49,969 individuals of European ancestry, 21,026 individuals of African ancestry and 14,517 individuals of Latino/Native American ancestry. We converted the highest education level attained to a binary trait based on those who attained a college degree versus those who did not and compared the frequency of *AFF3* expansions between the two groups. Associations were performed separately in each of the 12 sub-groups using Firth’s logistic regression^82^ due to the small sample sizes in some groups, incorporating the same covariates and options as utilized for the discovery PheWAS in UKB. Results for each sequencing center and ancestral group were then combined using z-score based meta-analyses with *METAL*.

### Identification of repeat expansions in The 100,000 Genomes Project

The 100,000 Genomes Project (100kGP) is a diagnostic and research study aimed at sequencing genomes from ∼70,000 patients seen within the UK National Health Service with either selected rare diseases or cancers. In the rare disease arm of the project, each proband was recruited under a specific disease and clinical data linked to each proband was collected including the clinical signs and symptoms encoded using human phenotype ontology (HPO). From this cohort, we selected 16,665 individuals: 7,871 probands with intellectual disability (defined as individuals whose clinical data included “intellectual disability” as an HPO term) and 8,794 unrelated controls (defined as probands not recruited under any neurological disease). Ancestry assignments for each individual were performed as described above using a set of high-quality LD-pruned SNVs. We performed genotyping of the *AFF3* TR in PCR-free 150bp paired-end Illumina GS data using ExpansionHunter (v3.2)^21^. We performed manual inspection of read alignment plots for all repeat alleles genotyped as >150bp and those deemed to be false positives due to poor read support or low-quality alignments over the TR were excluded. To avoid introduction of potential bias, manual classification of read alignment plots was performed by a user who was blind to the phenotype of the individual.

### Long read sequencing of trios with AFF3 expansions

DNA samples from two parent-offspring trios underwent HiFi sequencing using a Pacific Biosciences Revio instrument. Approximately 5 µg of genomic DNA were sheared to a target size of 15-20 kb using a Megaruptor 3 and library preparation was performed using the SMRTbell kit v3.0. Sequencing was performed on a Revio system using 24-hour movies. HiFi reads with methylation calls were generated on the Revio system and aligned to the GRCh38 assembly using pbmm2 (v1.9.0). We genotyped 937,122 TR loci with TRGT (v0.4.0) using default parameters and identified *de novo* TR mutations with TRGT-denovo (v0.1.0) using default parameters.

### Calculating polygenic risk score for educational attainment

We utilized data for 3,952 SNVs identified from GWAS^41^ to calculate a polygenic risk score (PRS) for educational attainment for each individual in the UKB. SNV genotypes were extracted from the vcf file derived from GS data, removing any genotypes with read depth <10 or quality score <20. For each of the 3,952 SNVs for which genotypes were available, at each SNV we multiplied the SNV effect size by the number of effect alleles present and summed these scores across all SNVs per individual to create a single numerical value per individual. The resulting PRS scores were then ranked and divided into quintiles, where quintile 1 represented the 20% of individuals with the highest PRS for educational attainment.

### Identification of recurrent microdeletions/microduplications

Using available GS data, we applied mosdepth to calculate the normalized read depth for each coding and untranslated region extended by ±100 bp per RefSeq gene, as described previously^76^. We selected six recurrent microdeletion/duplication regions^83^ that are known to have effects on neurodevelopment^84^ and occur at sufficient frequencies in the UKB cohort for robust statistical analyses (1q21.1, 15q11.2, 15q13.3, 16p13.11, 16p11.2 and 22q11.2). Individuals with deletions and duplications of these six regions were identified based on being clear outliers with an estimated copy number of ∼1 or ∼3 through visual inspection of the median read depth of all genes located within the CNV region (Supplementary Figure 19).

### Identification of SNVs that associate with the *AFF3* TRE

For the 168,641 unrelated individuals of European ancestry in the UKB utilized for PheWAS, SNV genotypes with MAF ≥0.001 located within ±500 kb of the *AFF3* TR were extracted from the vcf file derived from GS data, removing any genotypes with read depth <10 or quality score <20. We utilized REGENIE to perform association analysis between the 319 individuals we defined as carrying expansions of the GCC TR within *AFF3* at the Very Low threshold and 151,777 individuals in the bottom 90% of the population distribution by *AFF3* TR allele size, incorporating covariates of sex, age, GS insert size and the top five principle components derived from analysis of SNVs to account for ancestry. Associations were performed separately in samples sequenced by deCODE and Sanger Center, before combining results using z-score based meta-analyses in *METAL*. Results were then joined with SNVs from the GWAS Catalog with p<5x10^-8^ for the following traits: Educational attainment, Educational attainment (years of education), Educational attainment (college completion), Educational attainment (MTAG), Intelligence, Intelligence (MTAG), Cognitive ability, Cognitive ability (MTAG) and General cognitive ability.

## Data Availability

Information on how to access the data produced in the present work are contained within the manuscript.

https://ega-archive.org/studies/EGAS00001004587

https://www.ncbi.nlm.nih.gov/geo/query/acc.cgi?acc=GSE213478

https://storage.googleapis.com/gtex_analysis_v8/rna_seq_data/GTEx_Analysis_2017-06-05_v8_RNASeQCv1.1.9_gene_tpm.gct.gz

https://www.google.com/url?q=https://console.cloud.google.com/storage/browser/fc-secure-ff8156a3-ddf3-42e4-9211-0fd89da62108/GTEx_Analysis_2017-06-05_v8_ASE_WASP_counts_by_subject/&sa=D&source=docs&ust=1678389406863768&usg=AOvVaw1McwIkcmsHEds7G5NSMcf6

https://biobank.ctsu.ox.ac.uk/crystal/search.cgi

https://www.genomicsengland.co.uk/research/academic/join-gecip

https://github.com/bharatij/ExpansionHunter_Classifier

https://github.com/bharatij/Global-Ancestry-Assignment

https://www.researchallofus.org/data-tools/workbench/

## ACKNOWLEDGMENTS

We thank Prof. Karen Temple, University of Southampton, UK, for her support. This work was supported by NIH grants AG075051, NS105781, HD103782 and NS120241 to A.J.S., NHLBI Biodata Catalyst fellowship 5120339 to A.M.T., funding from The Prinses Beatrix Spierfonds (W.OR20-08) to J.J.F.A.V., funding from the European Research Council (ERC) under the European Union’s Horizon 2020 research and innovation programme (grant agreement 772376 – EScORIAL) to J.V. and funding from the UKRI (MR/S006753/1), Barts charity (MGU0569) and a Medical Research Council Clinician Scientist award (MR/S006753/1) to A.T.

This work was supported in part through the computational resources and staff expertise provided by Scientific Computing at the Icahn School of Medicine at Mount Sinai and supported by the Clinical and Translational Science Awards (CTSA) grant UL1TR004419 from the National Center for Advancing Translational Sciences. Research reported in this paper was supported by the Office of Research Infrastructure of the National Institutes of Health under award number S10OD026880. The content is solely the responsibility of the authors and does not necessarily represent the official views of the National Institutes of Health.

This research was made possible through access to the data and findings generated by the 100,000 Genomes Project. The 100,000 Genomes Project is managed by Genomics England Limited (a wholly owned company of the Department of Health and Social Care). The 100,000 Genomes Project is funded by the National Institute for Health Research and NHS England. The Wellcome Trust, Cancer Research UK and the Medical Research Council have also funded research infrastructure. The 100,000 Genomes Project uses data provided by patients and collected by the National Health Service as part of their care and support.

The All of Us Research Program is supported by the National Institutes of Health, Office of the Director: Regional Medical Centers: 1 OT2 OD026549; 1 OT2 OD026554; 1 OT2 OD026557; 1 OT2 OD026556; 1 OT2 OD026550; 1 OT2 OD 026552; 1 OT2 OD026553; 1 OT2 OD026548; 1 OT2 OD026551; 1 OT2 OD026555; IAA #: AOD 16037; Federally Qualified Health Centers: HHSN 263201600085U; Data and Research Center: 5 U2C OD023196; Biobank: 1 U24 OD023121; The Participant Center: U24 OD023176; Participant Technology Systems Center: 1 U24 OD023163; Communications and Engagement: 3 OT2 OD023205; 3 OT2 OD023206; and Community Partners: 1 OT2 OD025277; 3 OT2 OD025315; 1 OT2 OD025337; 1 OT2 OD025276. In addition, the All of Us Research Program would not be possible without the partnership of its participants.

## DATA AND CODE AVAILABILITY

Project MinE DNA methylation data are available from the European Genome-phenome Archive via accession EGAS00001004587

GTEx DNA methylation data, https://www.ncbi.nlm.nih.gov/geo/query/acc.cgi?acc=GSE213478

GTEx normalized expression level data, https://storage.googleapis.com/gtex_analysis_v8/rna_seq_data/GTEx_Analysis_2017-06-05_v8_RNASeQCv1.1.9_gene_tpm.gct.gz

GTEx allele specific expression data, https://www.google.com/url?q=https://console.cloud.google.com/storage/browser/fc-secure-ff8156a3-ddf3-42e4-9211-0fd89da62108/GTEx_Analysis_2017-06-05_v8_ASE_WASP_counts_by_subject/&sa=D&source=docs&ust=1678389406863768&usg=AOvVaw1McwIkcmsHEds7G5NSMcf6

UKB Data Showcase, https://biobank.ctsu.ox.ac.uk/crystal/search.cgi

Information on how to access the 100kGP data by joining a Genomics England Clinical Interpretation Partnership is available online, www.genomicsengland.co.uk/join-a-gecip-domain

All of Us Researcher Workbench, https://www.researchallofus.org/data-tools/workbench/

Code utilized for this study is available at GitHub, as follows:

https://github.com/bharatij/ExpansionHunter_Classifier

https://github.com/bharatij/Global-Ancestry-Assignment

https://github.com/PacificBiosciences/pbmm2

https://github.com/PacificBiosciences/trgt

https://github.com/PacificBiosciences/trgt-denovo

https://zenodo.org/records/7987365#.ZHY9TOzMJAc

## Author contributions

B.J., P.G., J.J.F.A.V., K.I., W.L. T.M. and E.D. designed bioinformatics pipelines and performed data analyses. D.G., M.J., A.M.T., S.L.G., C.R. and M.B. performed data analyses. N.L. and K.L. contributed clinical information. H.H., B.P., J.V. and A.T. supervised and advised on the project. A.J.S. conceived the study, performed data analyses, supervised the project and drafted the manuscript. All authors reviewed and approved the final draft.

