## Supplemental Figures 1-23 for "A phenome-wide association study of methylated GC-rich repeats identifies a GCC repeat expansion in *AFF3* as a significant cause of intellectual disability"

**Supplementary Information**

**The Genomics England Research Consortium**

Ambrose, J. C. ^1^; Arumugam, P. ^1^; Bevers, R. ^1^; Bleda, M. ^1^; Boardman-Pretty, F. ^1,2^; Boustred, C. R. ^1^; Brittain, H. ^1^; B wn, M.A.; Caulfield, M. J. ^1,2^; Chan, G. C. ^1^; Giess A. ^1^; Griffin, J. N. ^1^; Hamblin, A. ^1^; Henderson, S. ^1,2^; Hubbard, T. J. P. ^1^; Jackson, R. ^1^; Jones, L. J. ^1,2^; Kasperaviciute, D. ^1,2^; Kayikci, M. ^1^; Kousathanas, A. ^1^; Lahnstein, L. ^1^; Lakey, A. ^1;^ Leigh, S. E. A. ^1^; Leong, I. U. S. ^1^; Lopez, F. J. ^1^; Maleady-Crowe, F. ^1^; McEntagart, M. ^1^; Minneci F. ^1^; Mitchell, J. ^1^; Moutsianas, L. ^1,2^; Mueller, M. ^1,2^; Murugaesu, N. ^1^; Need, A. C. ^1,2^; O‘Donovan P. ^1^; Odhams, C. A. ^1^; Patch, C. ^1,2^; Perez-Gil, D. ^1^; Pereira, M. B. ^1^; Pullinger, J. ^1^; Rahim, T. ^1^; Rendon, A. ^1^; Rogers, T. ^1^; Savage, K. ^1^; Sawant, K. ^1^; Scott, R. H. ^1^; Siddiq, A. ^1^; Sieghart, A. ^1^; Smith, S. C. ^1^; Sosinsky, A. ^1,2^; Stuckey, A. ^1^; Tanguy M. ^1^; Taylor Tavares, A. L. ^1^; Thomas, E. R. A. ^1,2^; Thompson, S. R. ^1^; Tucci, A. ^1,2^; Welland, M. J. ^1^; Williams, E. ^1^; Witkowska, K. ^1,2^; Wood, S. M. ^1,2^; Zarowiecki, M. ^1^

*^1^ Genomics England, London, UK*

*^2^ William Harvey Research Institute, Queen Mary University of London, London, EC1M 6BQ, UK*

**Project MinE ALS Sequencing Consortium**

Al-Chalabi, A. ^1^; Andersen, P. ^2^; Başak, N. A. ^3^; Berg, L. H. ^4^; Carvalho, M. ^5,6^; Corcia, P. ^7,8^; Couratier, P. ^7^; Dalgard, C. L. ^9^; Damme, P. ^10,11^; Drory, V. ^12^; Glass, J. D. ^13,14^; Gotkine, M. ^15^; Hardiman, O. ^16,17^; Landers, J. E. ^18^; McLaughin, R. ^19^; Mora Pardina, J. S. ^20^; Morrison, K. E. ^21^; Povedano, M. ^22^; Shaw, C. ^1^; Shaw, P. J. ^23^; Silani, V. ^24,25^; Ticozzi, N. ^24,25^; Veldink, J. H. ^4^; Vourc'h, P. ^26,27^; Weber, M. ^28^

*^1^ Maurice Wohl Clinical Neuroscience Institute, King's College London, Department of Basic and Clinical Neuroscience, London, UK*

*^2^ Department of Clinical Science, Neurosciences, Umeå University, Sweden*

*^3^ Koç University, School of Medicine, KUTTAM-NDAL, Istanbul Turkey*

*^4^ Department of Neurology, UMC Utrecht Brain Center, University Medical Center Utrecht, Utrecht University, Utrecht, The Netherlands*

*^5^ Instituto de Fisiologia, Instituto de Medicina Molecular,Faculdade de Medicina, Universidade de Lisboa, Lisbon, Portugal*

*^6^ Department of Neurosciences, Hospital de Santa Maria-CHLN, Lisbon, Portugal*

*^7^ Centre SLA, CHRU de Tours, Tours, France*

*^8^ UMR 1253, iBrain, Université de Tours, Inserm, Tours, France*

*^9^ The American Genome Center, Uniformed Services University - "America's Medical School", Bethesda, MD, USA*

*^10^ KU Leuven - University of Leuven, Department of Neurosciences*

*^11^ VIB, Center for Brain & Disease Research, Laboratory of Neurobiology, Leuven, Belgium*

*^12^ Department of Neurology Tel-Aviv Sourasky Medical Centre , Israel*

*^13^ Department Neurology, Emory University School of Medicine, Atlanta, GA, USA*

*^14^ Emory ALS Center, Emory University School of Medicine, Atlanta, GA, USA*

*^15^ Department of Neurology, Hadassah Medical Organization and Faculty of Medicine, Hebrew University of Jerusalem, Israel*

*^16^ Academic Unit of Neurology, Trinity College Dublin, Trinity Biomedical Sciences Institute, Dublin, Republic of Ireland*

*^17^ Department of Neurology, Beaumont Hospital, Dublin, Republic of Ireland*

*^18^ Department of Neurology, University of Massachusetts Medical School, Worcester, MA, USA*

*^19^ Complex Trait Genomics Laboratory, Smurfit Institute of Genetics, Trinity College Dublin, Dublin, Republic of Ireland*

*^20^ ALS Unit, Hospital San Rafael, Madrid, Spain*

*^21^ School of Medicine, Dentistry and Biomedical Sciences, Queen’s University Belfast, UK*

*^22^ la Unitat Funcional de Motoneurona, Cap de Secció de Neurofisiologia, Servei de Neurologia, Hospital Universitario de Bellvitge-IDIBELL, Spain*

*^23^ Sheffield Institute for Translational Neuroscience (SITraN), University of Sheffield, Sheffield, UK*

*^24^ Department of Neurology and Laboratory of Neuroscience, IRCCS Istituto Auxologico Italiano, Milano, Italy*

*^25^ Department of Pathophysiology and Tranplantation, ‘Dino Ferrari’ Center, Università degli Studi di Milano, Milano, Italy*

*^26^ Service de Biochimie et Biologie moléculaire, CHU de Tours, Tours, France*

*^27^ UMR 1253, Université de Tours, Inserm, 37044 Tours, France*

*^28^ Neuromuscular Diseases Unit/ALS Clinic, Kantonsspital St. Gallen, 9007, St. Gallen, Switzerland*

**Supplementary Tables**

**Supplementary Table 1.** DMRs found in MinE samples with the 450k array.

**Supplementary Table 2.** DMRs found in MinE samples with the 850k array.

**Supplementary Table 3.** Coordinates of 28 GC-rich repeats genotyped with ExpansionHunter.

**Supplementary Table 4.** TRE calling thresholds.

**Supplementary Table 5.** TR allele sizes and number of expansions per threshold identified in the UK Biobank.

**Supplementary Table 6.** Results of PheWAS for 25 autosomal GC-rich repeats, showing all associations with p<0.001.

**Supplementary Table 7.** Results of PheWAS for 3 chrX GC-rich repeats, showing all associations with p<0.001.

**Supplementary Table 8.** Replication of *AFF3* expansions with reduced educational attainment in the All of Us cohort.

**Supplementary Table 9.** Ancestry estimates for individuals in the UK100k cohort with TREs of *AFF3*.

**Supplementary Table 10.** Recurrent phenotypic features reported in two or more probands with *AFF3* expansions in the 100kGP cohort.

**Supplementary Table 11.** Results of analysis with TRGT-denovo in two trios from the 100kGP cohort with *AFF3* expansions.

**Supplementary Table 12.** SNVs associated with the presence of the *AFF3* repeat expansion in the UKB cohort.

**Supplementary Table 13.** Definitions of 28 GC-rich repeats for genotyping with ExpansionHunter.

**Supplementary Table 14.** Validation rates for TRE calls before and after quality control with a Random Forest classifier using 1,027 All of Us samples sequenced using both Illumina and PacBio HiFi GS.

**Supplementary Table 15.** 10,615 UKB phenotypes tested showing the number of individuals with data for each trait.

**Supplementary Figures**

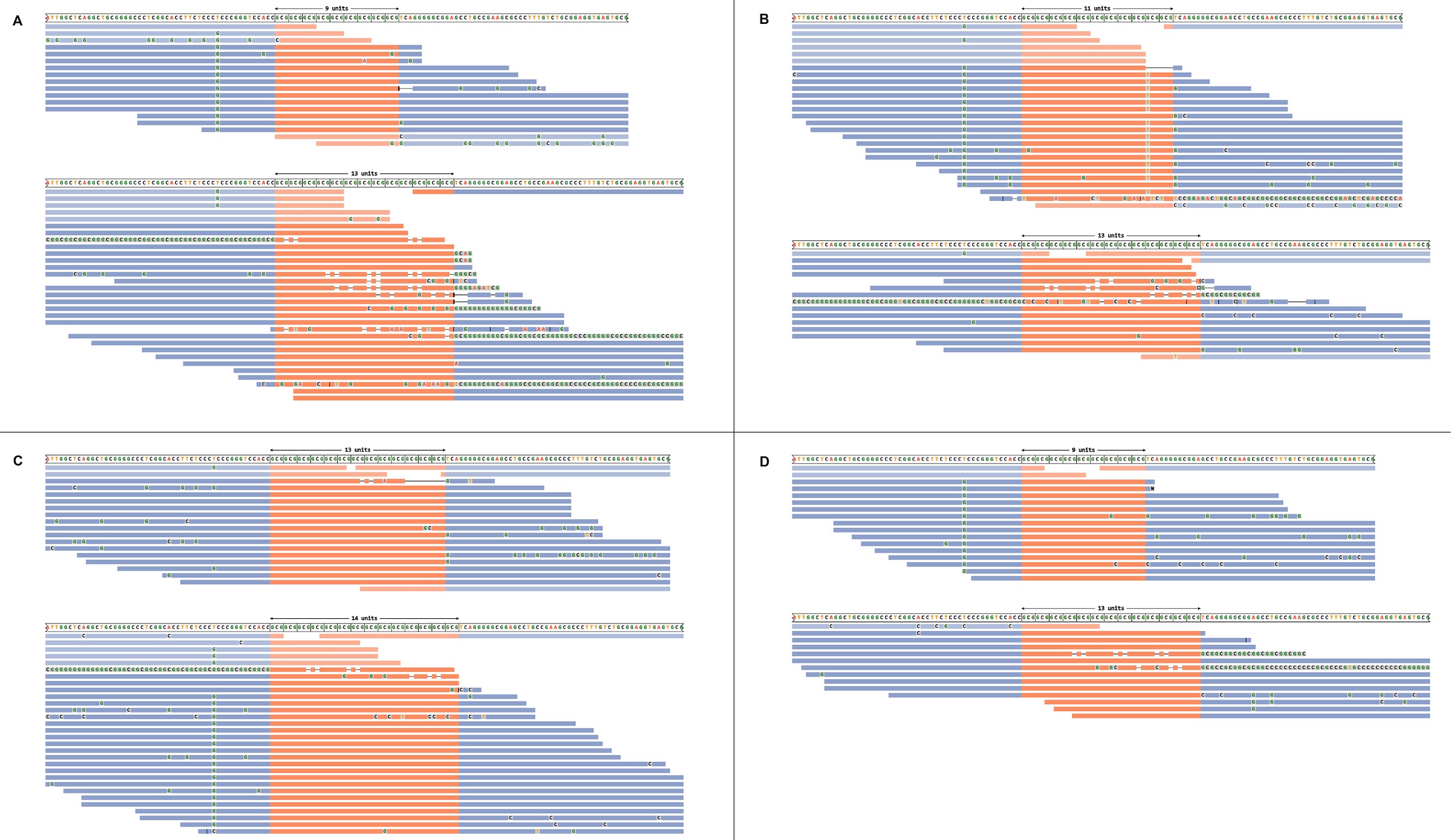

**Supplementary Figure 1. Potential false-negative TREs missed by ExpansionHunter** (**A-D**) Read alignment plots of the GCG TR at *ZNF713* generated using REViewer^1^ in four individuals with hypermethylation of *ZNF713*, but who were all genotyped by ExpansionHunter as having two TR alleles within the normal range. In all four individuals, one or more reads containing high TR copy number can be seen aligned to the region, suggesting the possible presence of a TRE that was not formally identified by ExpansionHunter. In each individual, alignments are shown grouped for each of the two alleles present.

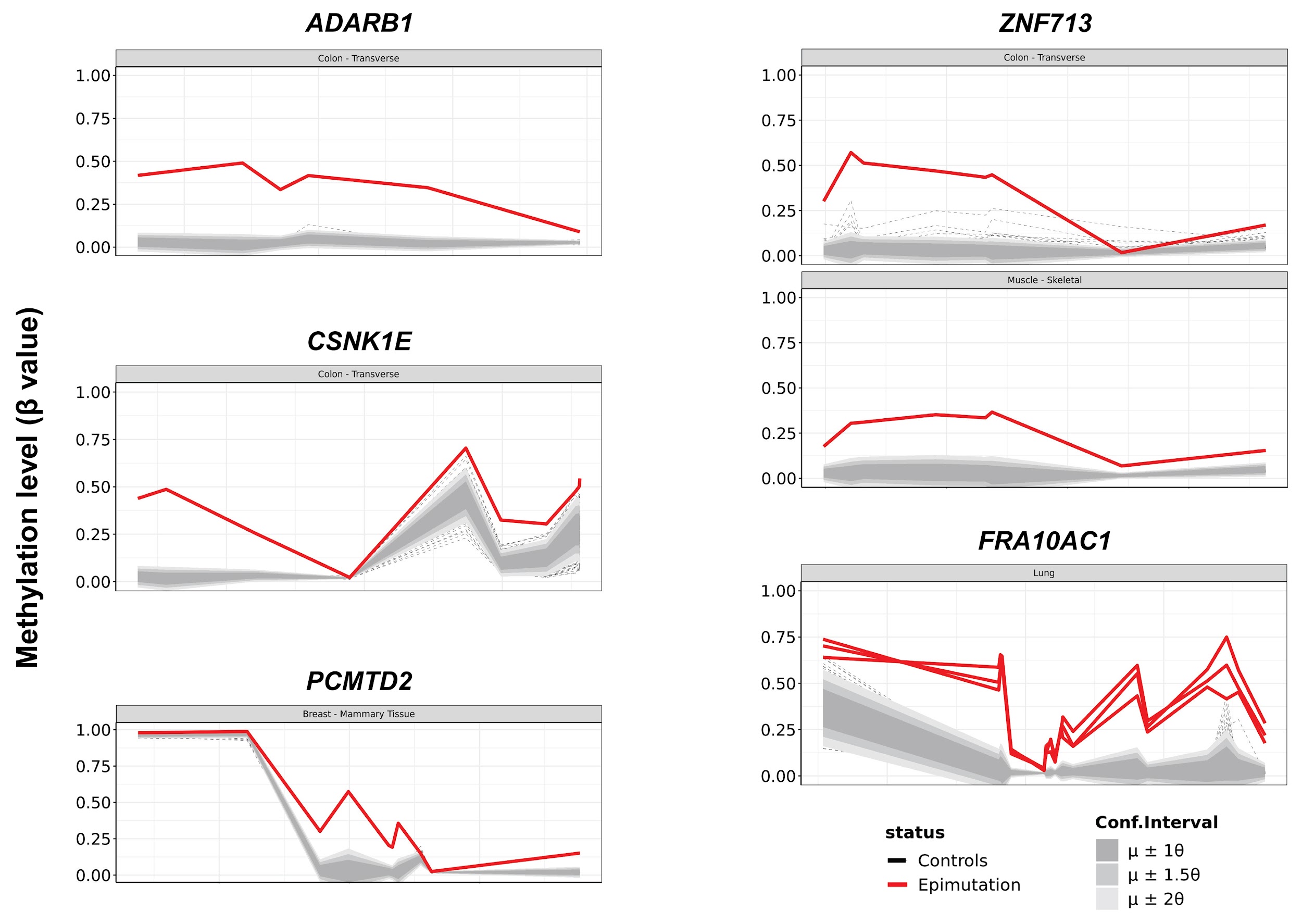

**Supplementary Figure 2. Methylated TREs identified in the GTEx cohort.** Utilizing our pipeline for identifying differentially methylated regions, we identified multiple individuals in the GTEx cohort that showed hypermethylation at some of the same loci found to be associated with GC-rich TREs, indicating that these individuals likely have large, methylated expansions of the underlying TR^2^. Individuals with hypermethylated DMRs are shown in red, while the distribution of methylation profiles in all other individuals profiled in that tissue are indicated by shades of gray, with methylation values lying outside ±2 standard deviations from the mean shown as dashed gray lines. Due to all individuals having undergone Illumina GS that utilized PCR during library preparation, we were unable to formally confirm the presence of these TREs in these individuals.

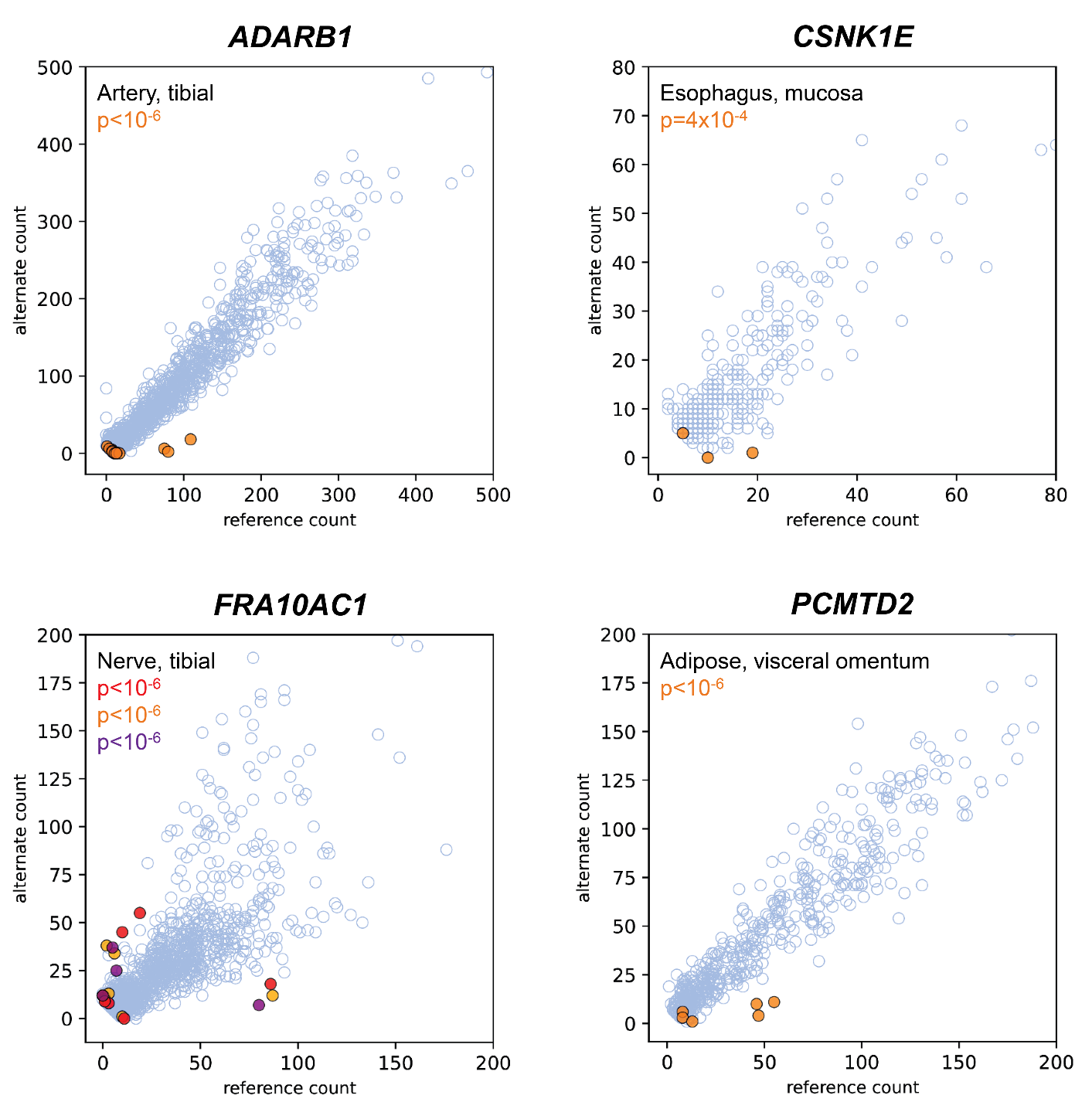

**Supplementary Figure 3. Hypermethylated TREs are associated with significant allelic expression bias in the GTEx cohort.** We analyzed available allelic expression data from individuals with hypermethylation of *ADARB1*, *CSNK1E*, *FRA10AC1* and *PCMTD2*. In each case, carriers of the presumptive TRE showed highly biased allelic expression, consistent with silencing in cis of the expanded/hypermethylated allele, as has been shown previously for other GC-rich TREs^3-6^. Each plot shows data from a selected tissue that had high mean expression level and a high number of available samples with informative heterozygous SNVs, including the TRE carrier. However, similar results were obtained in other tissues. Each data point represents a heterozygous SNV in one individual that had ≥10 overlapping RNAseq reads. Where an individual had >1 expressed heterozygous SNV in the gene, each is plotted as a separate point. Data from individuals with a methylated presumptive TRE of the gene are shown in orange, except for *FRA10AC1* where three different carrier individuals are shown (displayed in orange, red and purple, respectively), while data from all other available individuals from that same tissue are shown in blue. For carriers of other presumptive TREs, due to either low expression of the gene and/or few or no transcribed SNVs in the relevant individual, data were too sparse to draw robust conclusions on allelic expression bias.
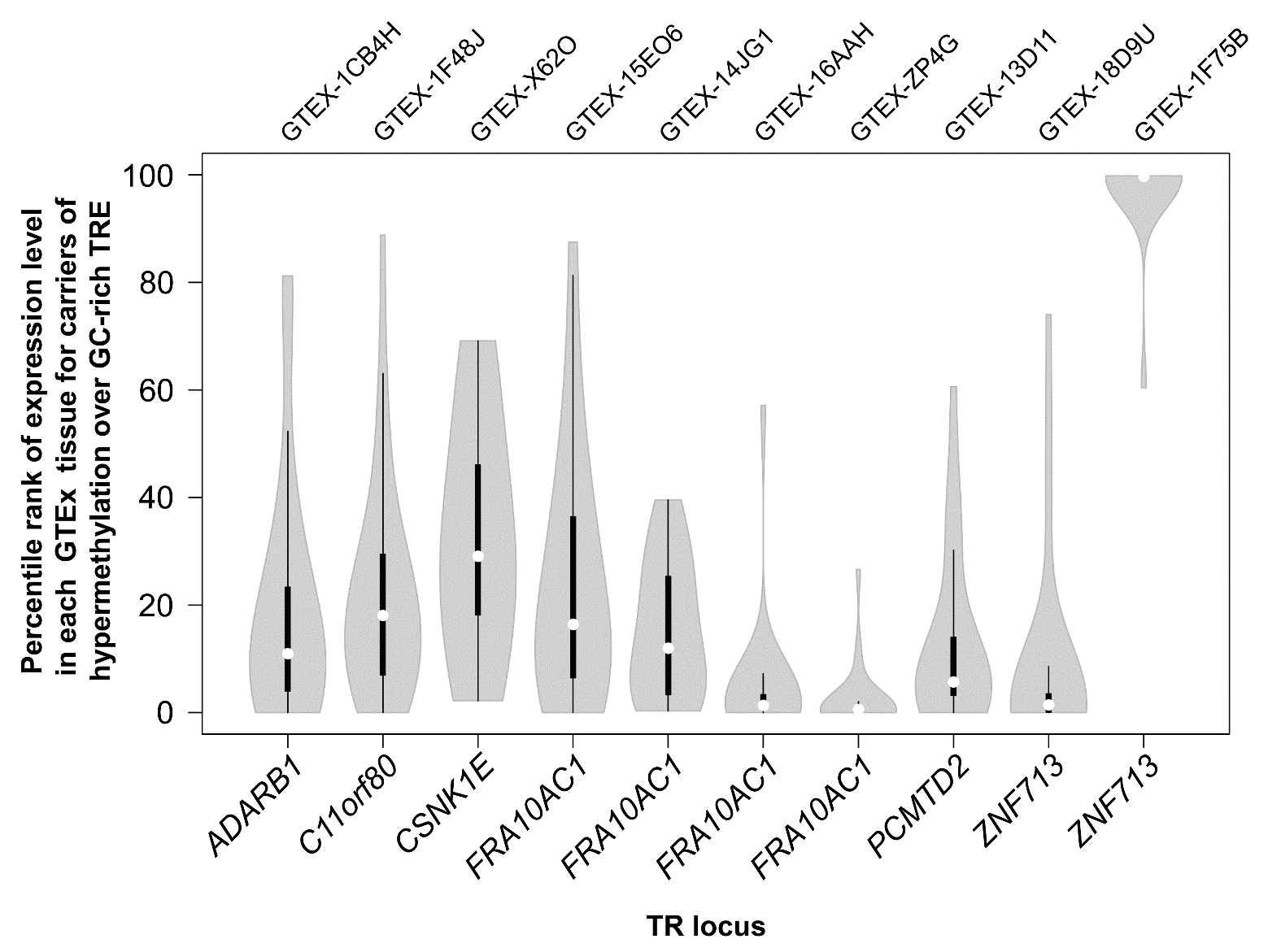
**Supplementary Figure 4. Hypermethylated TREs are associated with altered gene expression levels in multiple tissues of the GTEx cohort.** Using DNA methylation data, we identified putative carriers of methylated TREs in GTEx samples. In all cases except one, we observed that the individual carrying the presumptive TRE showed unusually low expression level of the associated gene, consistent with reduced or silenced expression of the methylated TRE allele in *cis*. In one individual with increased methylation of the *ZNF713* locus, we observed the opposite trend, with this individual consistently showing the highest *ZNF713* expression across multiple tissues. The methylation gain in this individual was lower than that observed in the other *ZNF713* epimutation carrier and we hypothesize this may indicate the presence of mosaicism for unmethylated premutation and methylated full mutation alleles, with the premutation showing unusually high expression level, as occurs for premutations of *FMR1*^3^ and *DIP2B^7^*. Plot shows the rank of expression level converted to a percentile in all tissues available for the individual carrying each presumptive TRE, with each violin showing data for a single TRE carrier in a mean of 21 different tissues per individual. In every case except for the carrier of a hypermethylated *CSNK1E* allele, permutation testing showed that the observed expression ranks in the individuals carrying the hypermethylated alleles across were significantly different from the null (p<10^-6^). Within each violin, white circles show the medians; box limits indicate the 25^th^ and 75^th^ percentiles; whiskers extend 1.5 times the interquartile range from the 25^th^ and 75^th^ percentiles. GTEx sample IDs are shown above the plot.

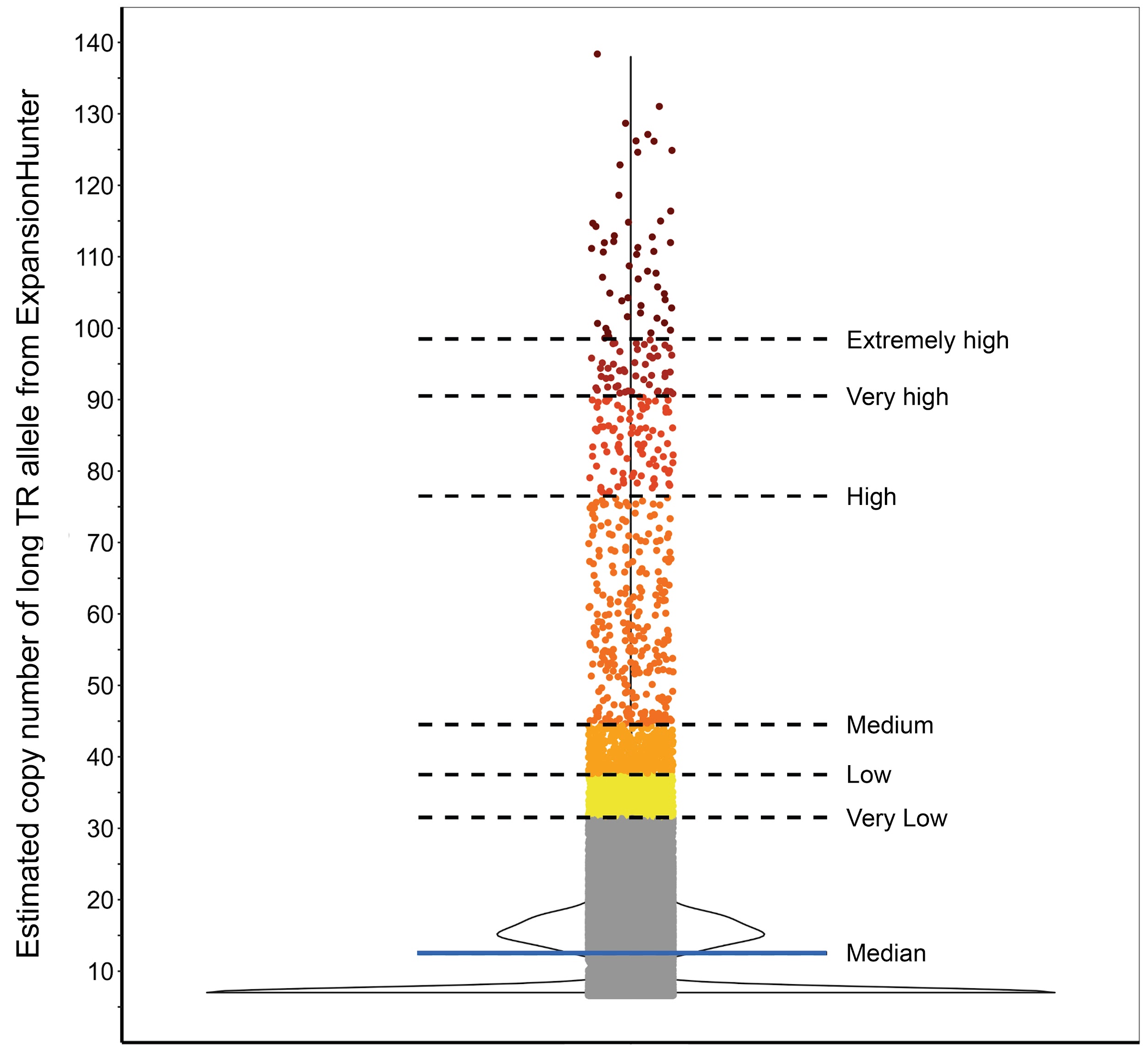

**Supplementary Figure 5.** Example plot showing how TREs were defined from TR genotypes in the population using six different outlier thresholds, as per the criteria listed in Supplementary Table 4. The plot shows data for *DIP2B* in the Sanger Center sub-cohort, with the violin showing data density and the median indicated by the blue horizontal line.

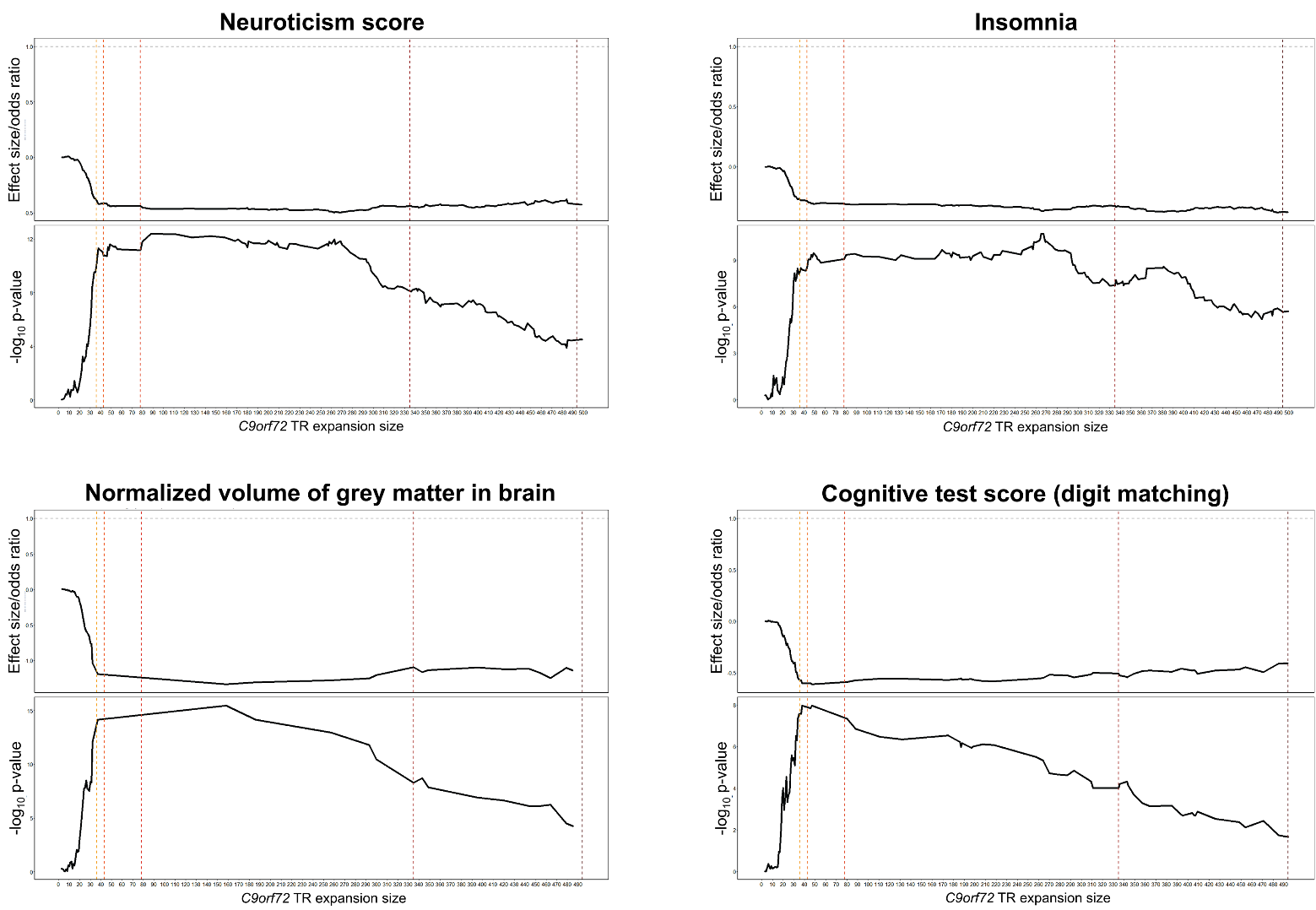

**Supplementary Figure 6. Invariant relationship between expansion size and effect size of four traits identified as significantly associated with the *C9orf72* expansion.** In each case, we observed that once the pathogenic threshold (≥30 copies) is reached, the effect size remains almost constant. Thus, we did not observe any evidence to support milder ALS symptoms at smaller *C9orf72* repeat sizes. For each trait, the upper panel shows odds ratio (for binary traits) or effect size (for quantitative traits), while the lower panel shows -log_10_ p-value of the association. Colored vertical dashed lines indicate the different thresholds used to call expanded alleles.

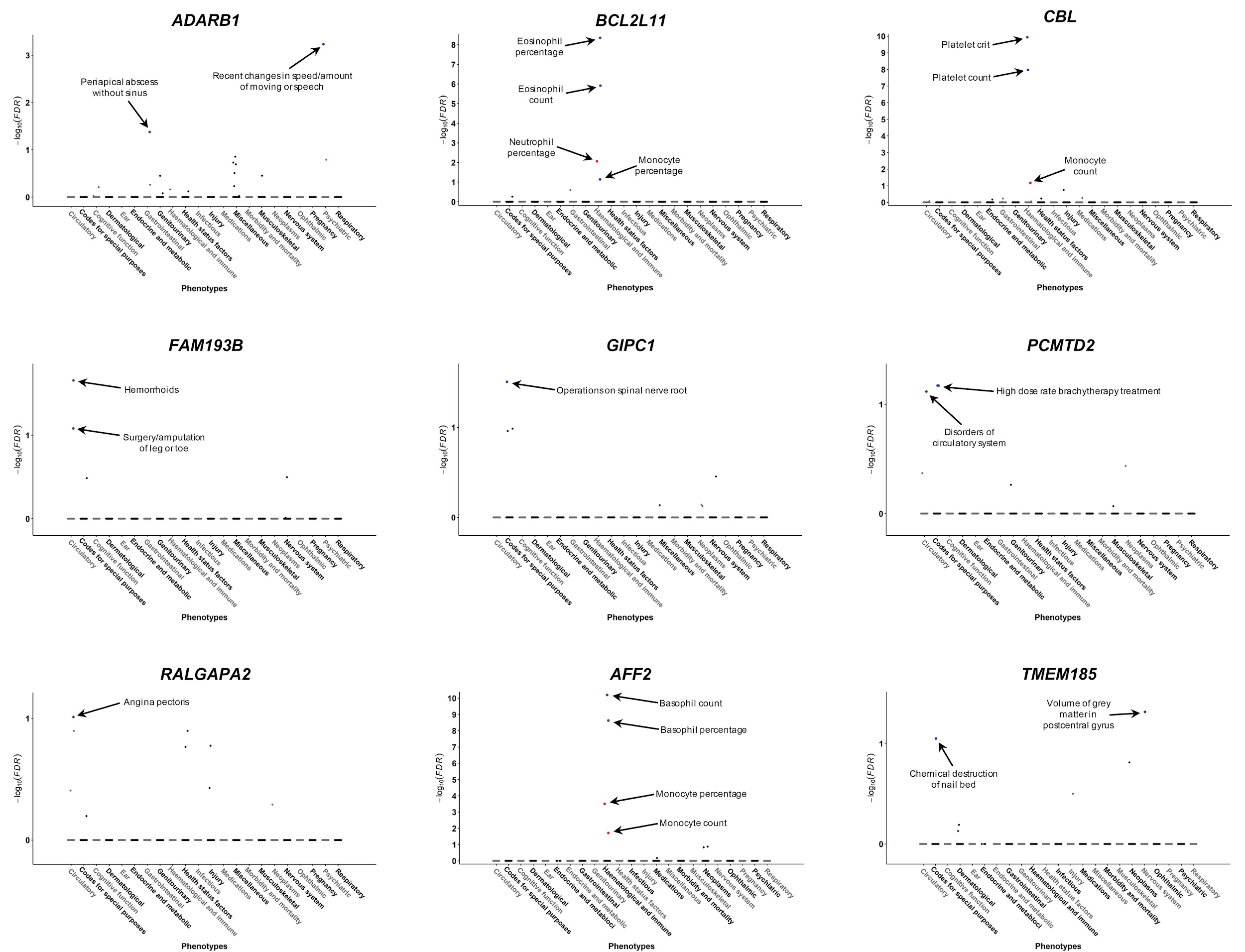

**Supplementary Figure 7. Results of phenome-wide association analysis for expansions of nine GC-rich tandem repeats.** Traits are grouped into physiological categories (x-axis) and we plot the -log_10_ FDR-corrected q-value (y-axis). Significant associations (FDR q<0.1) are individually labeled and shown in color to indicate directionality of effect (blue for positive associations, red for negative associations). Note that in some cases the trait names shown have been modified for clarity/brevity. Where a TRE:trait pair was significant under multiple definitions for identifying TREs (see Methods), we show only the most significant result for that TRE:trait pair.

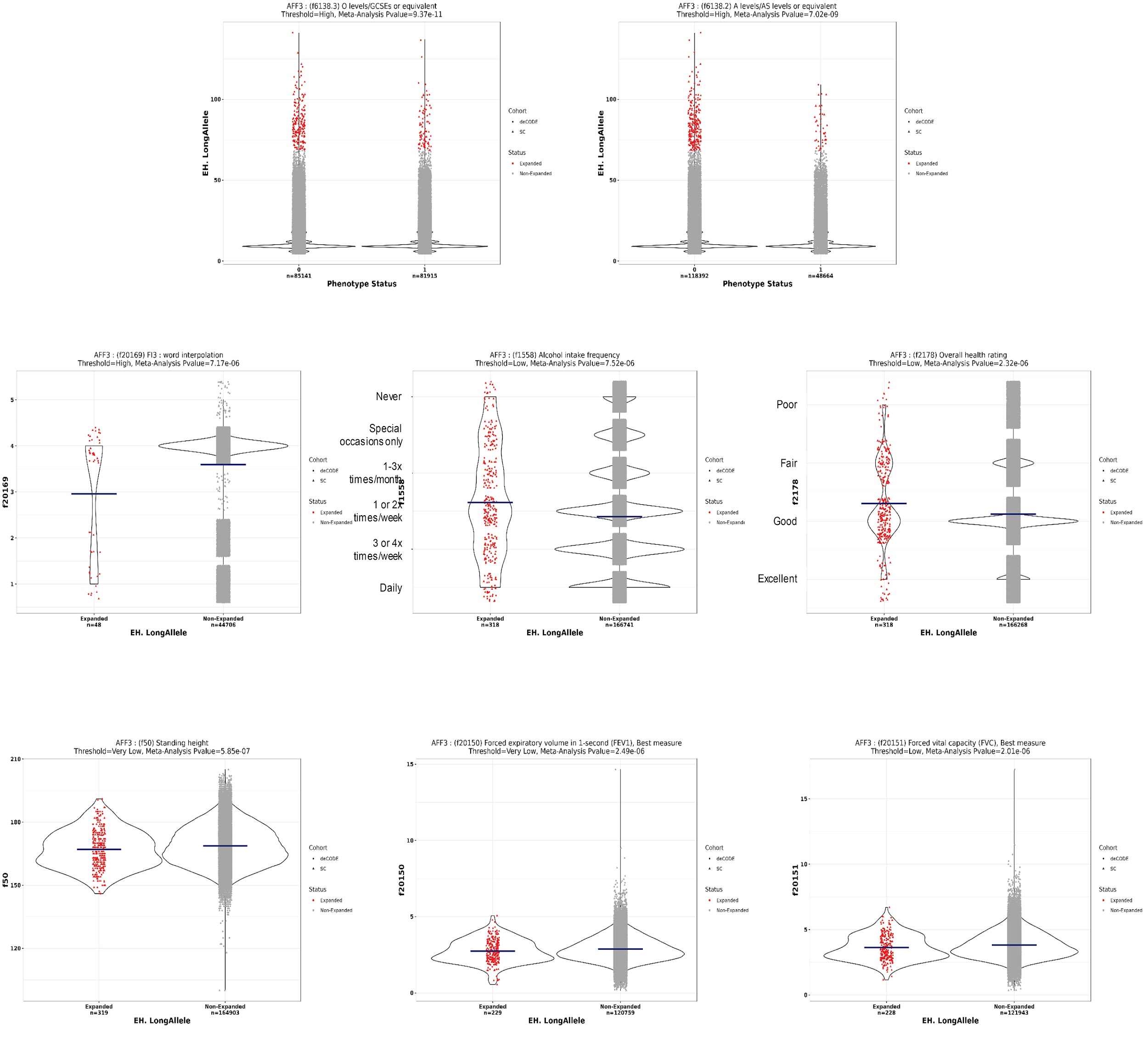

**Supplementary Figure 8.** Plots showing raw data for significant associations identified by PheWAS for expansions of *AFF3* in the UKB. Blue horizontal lines show the mean allele size of each distribution.

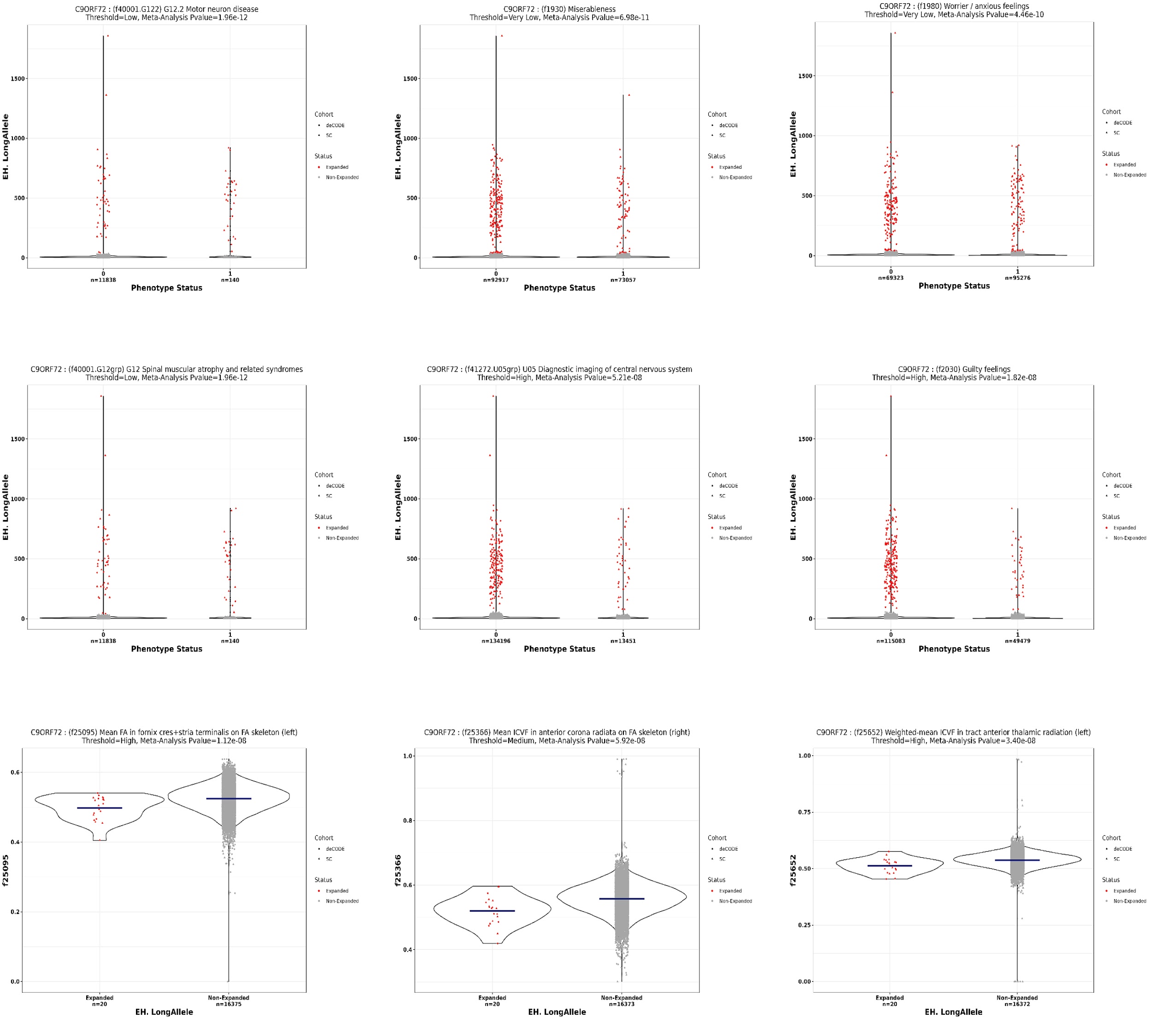

**Supplementary Figure 9.** Plots showing raw data for significant associations identified by PheWAS for expansions of *C9orf72* in the UKB. Blue horizontal lines show the mean allele size of each distribution.

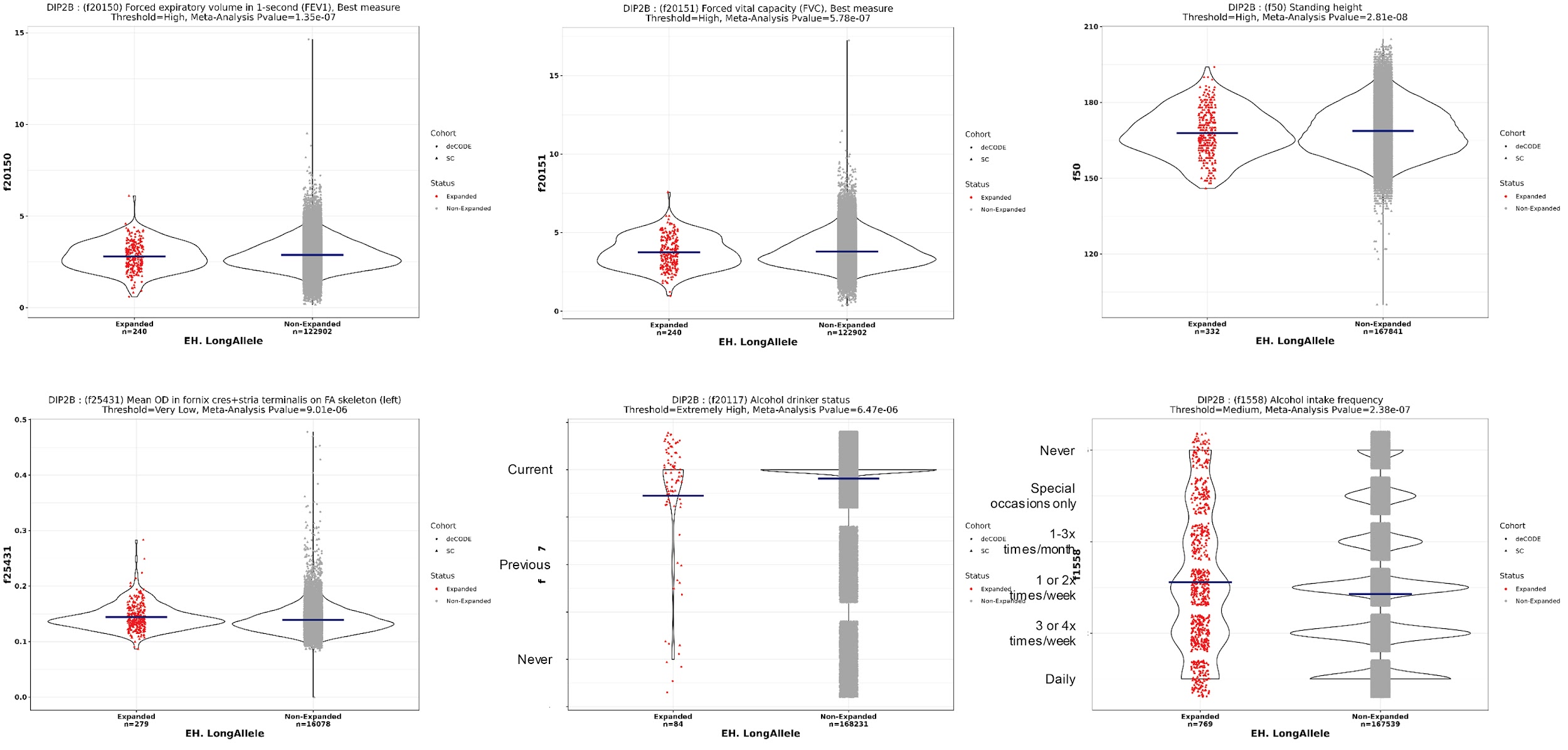

**Supplementary Figure 10.** Plots showing raw data for significant associations identified by PheWAS for expansions of *DIP2B* in the UKB. Blue horizontal lines show the mean allele size of each distribution.

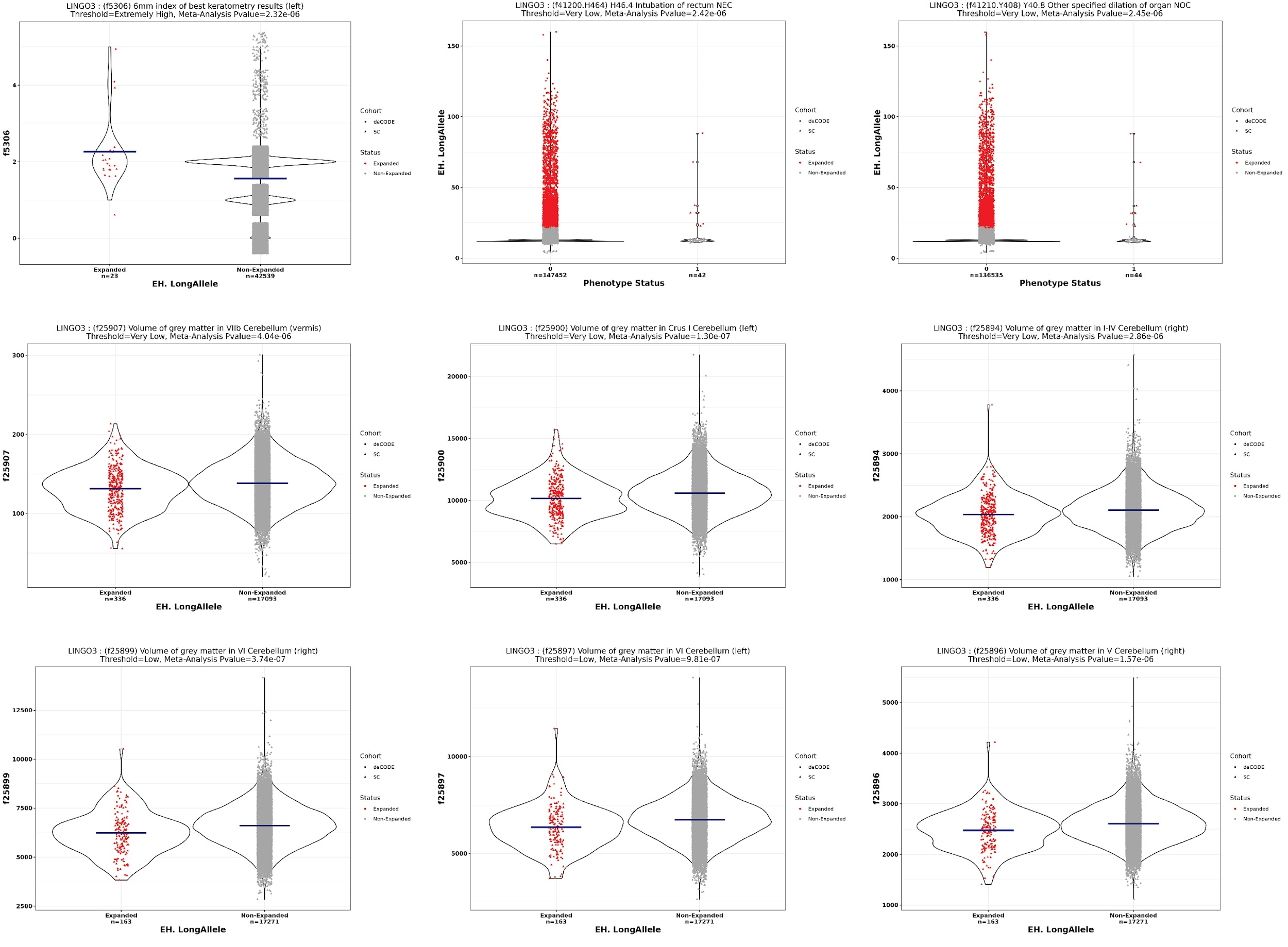

**Supplementary Figure 11.** Plots showing raw data for significant associations identified by PheWAS for expansions of *LINGO3* in the UKB. Blue horizontal lines show the mean allele size of each distribution.

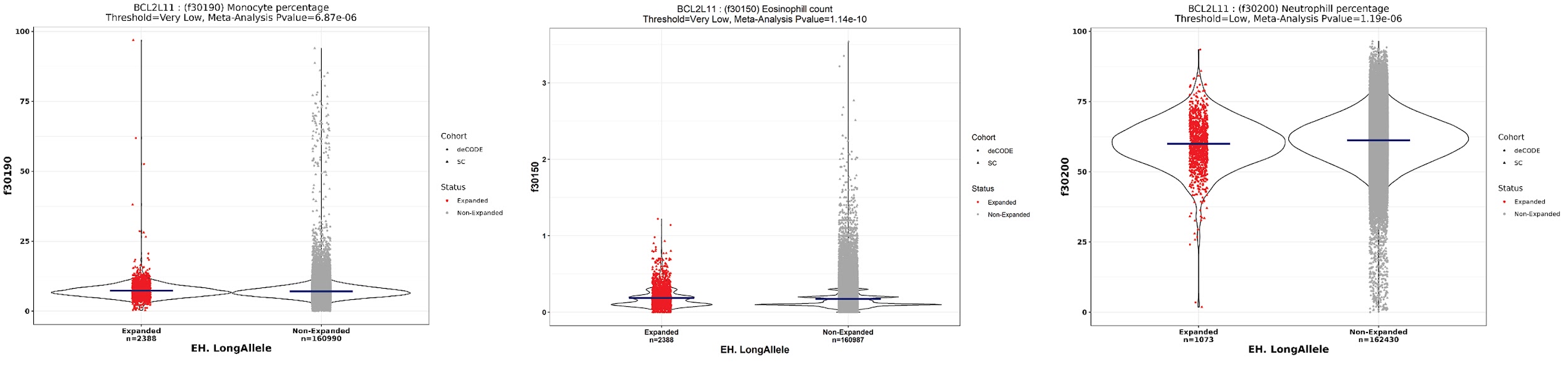

**Supplementary Figure 12.** Plots showing raw data for significant associations identified by PheWAS for expansions of *BCL2L11* in the UKB. Blue horizontal lines show the mean allele size of each distribution.

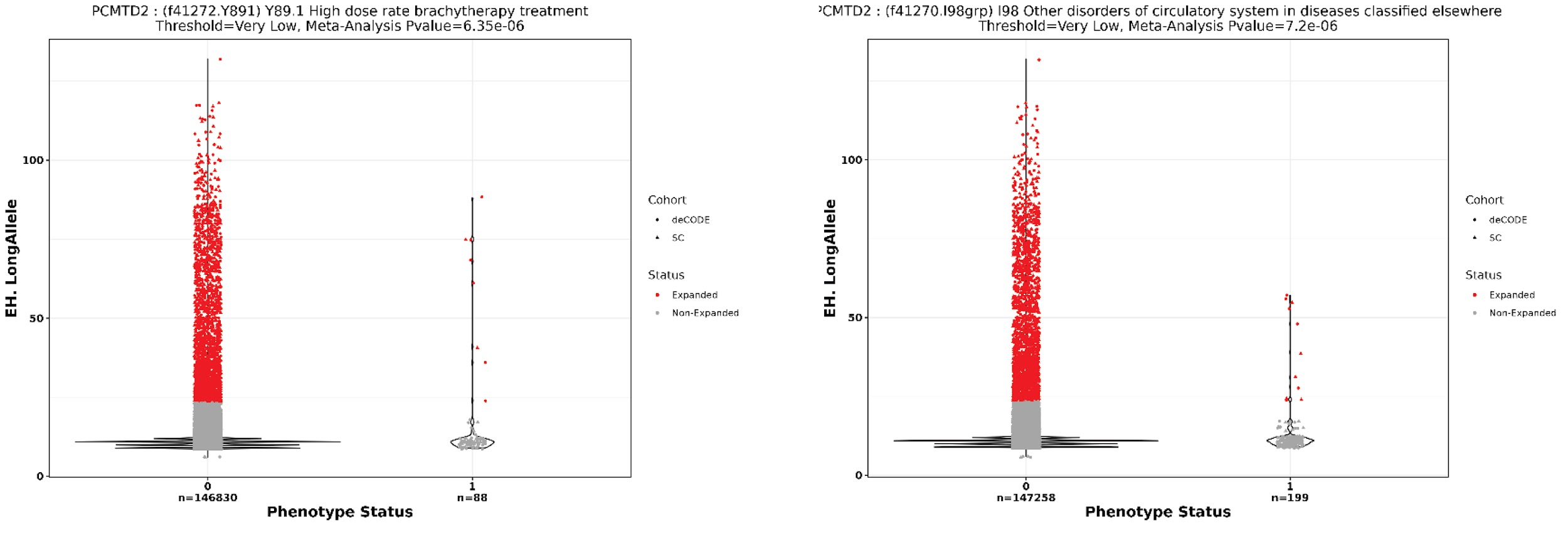

**Supplementary Figure 13.** Plots showing raw data for significant associations identified by PheWAS for expansions of *PCMTD2* in the UKB. Blue horizontal lines show the mean allele size of each distribution.

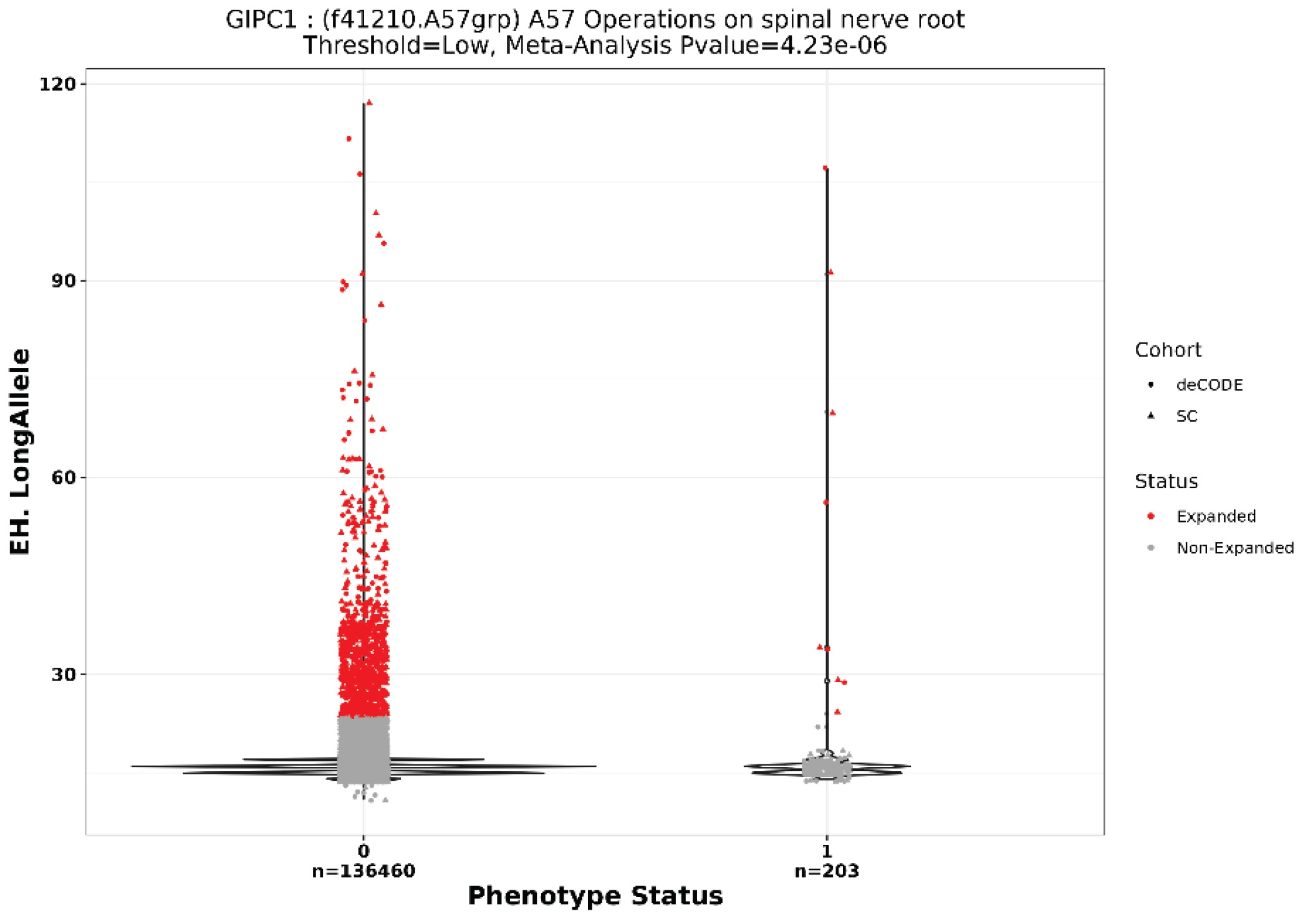

**Supplementary Figure 14.** Plot showing raw data for significant association identified by PheWAS for expansions of *GIPC1* in the UKB. Blue horizontal lines show the mean allele size of each distribution.

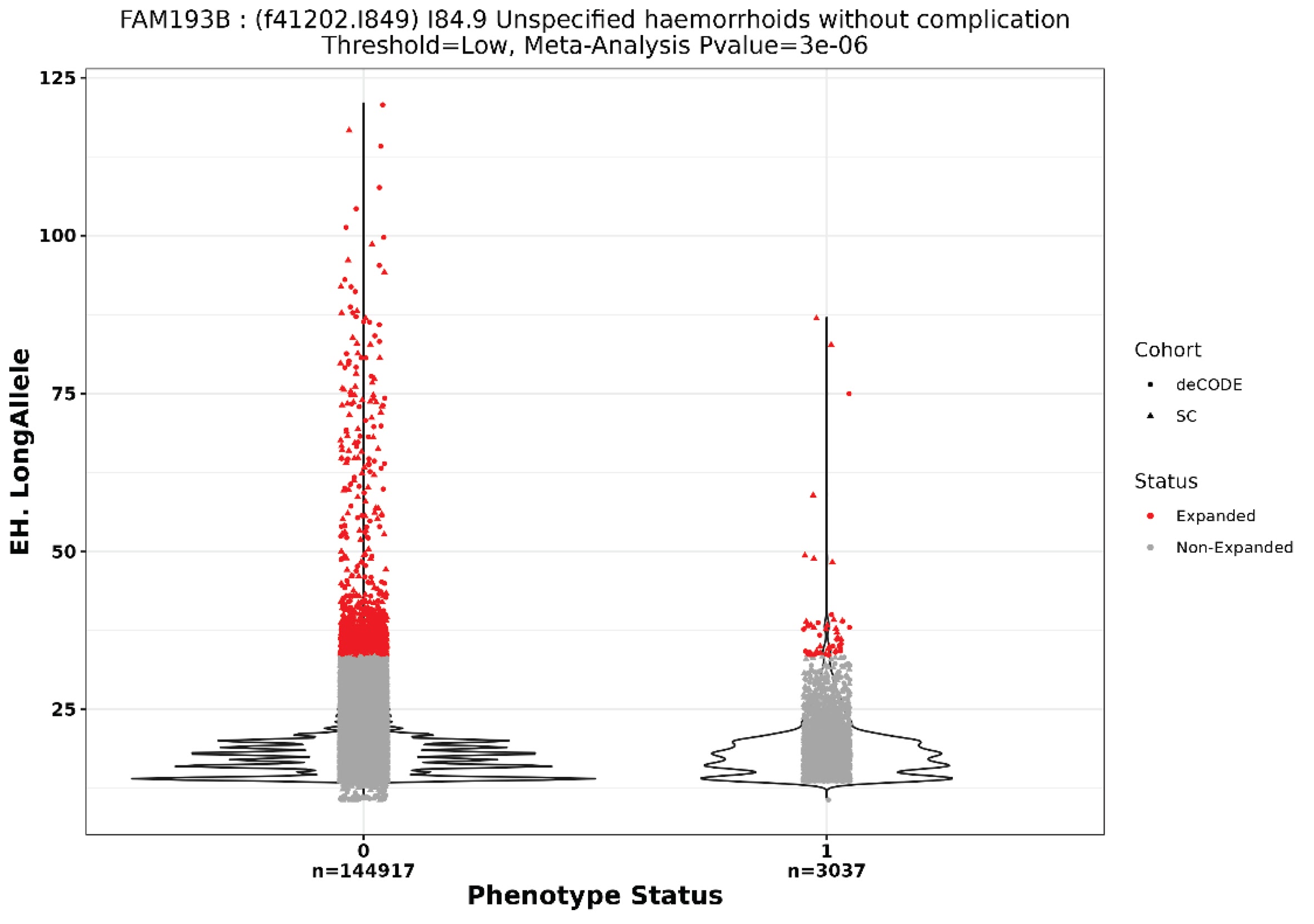

**Supplementary Figure 15.** Plot showing raw data for significant association identified by PheWAS for expansions of *FAM193B* in the UKB. Blue horizontal lines show the mean allele size of each distribution.

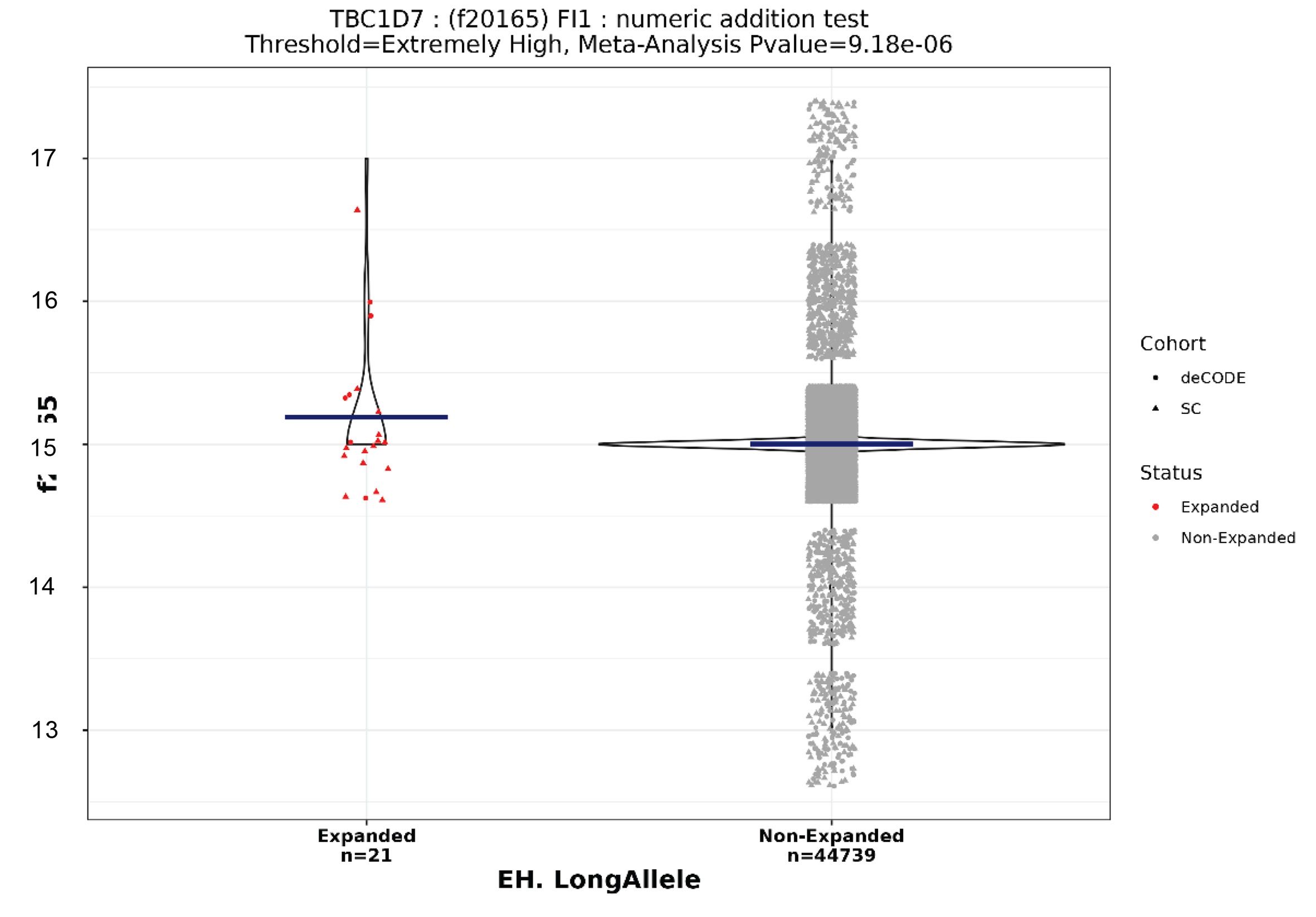

**Supplementary Figure 16.** Plot showing raw data for significant association identified by PheWAS for expansions of *TBC1D7* in the UKB. Blue horizontal lines show the mean allele size of each distribution.

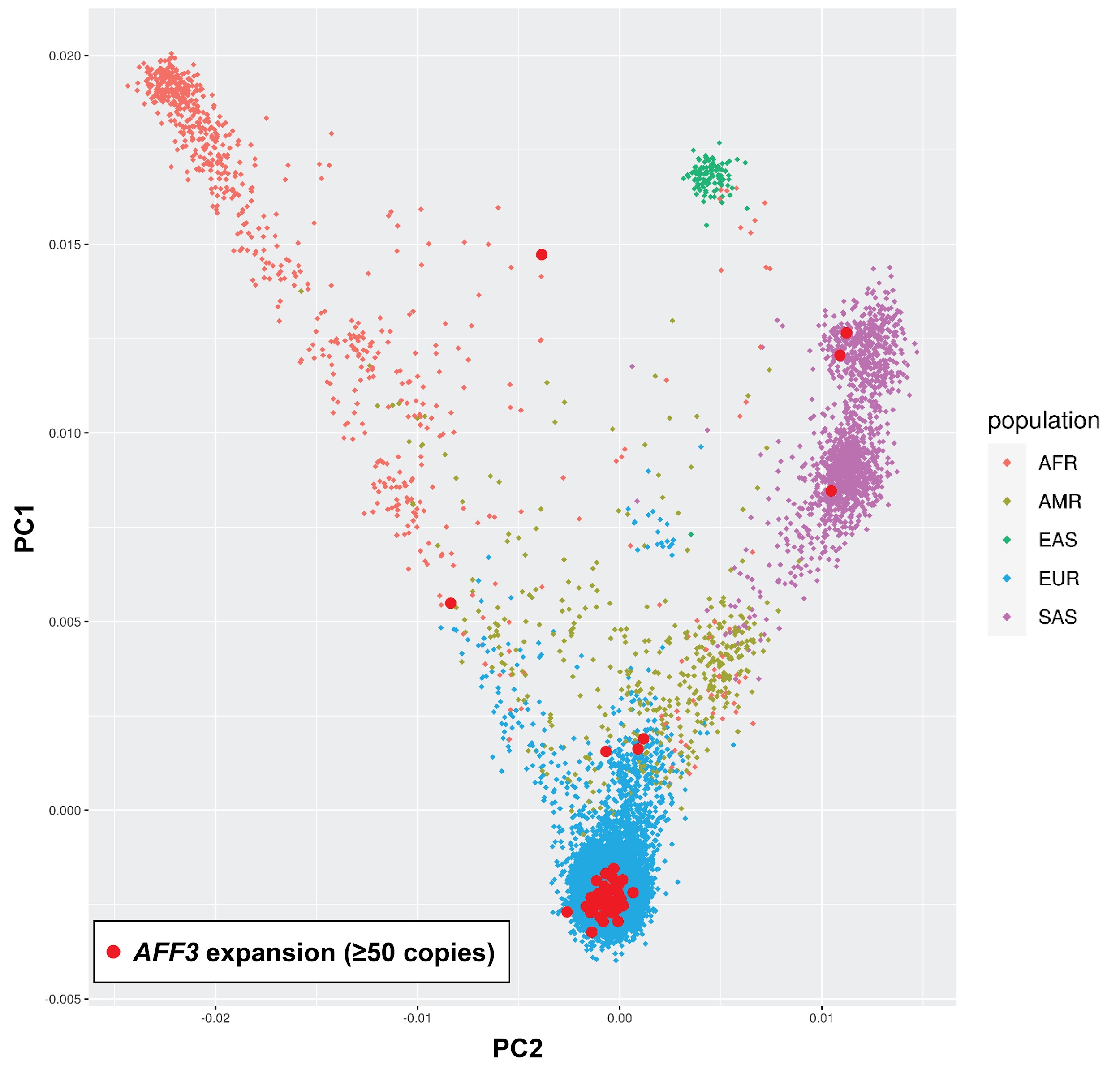

**Supplementary Figure 17.** PCA plot based on LD-pruned SNVs showing global ancestry estimates of individuals with *AFF3* TREs (red filled circles) compared to all other probands with unsolved ID and non-neurological controls in the 100kGP cohort. Overall, there were no significant differences in the frequencies of the *AFF3* TRE among ancestries that would confound the observed enrichment in unsolved probands with ID. For the purposes of this plot, *AFF3* TREs are defined as those genotyped by ExpansionHunter (v3.2) as ≥50 copies. A more detailed breakdown by global ancestry estimate, case:control status and *AFF3* TR size is shown in Supplementary Table 9.

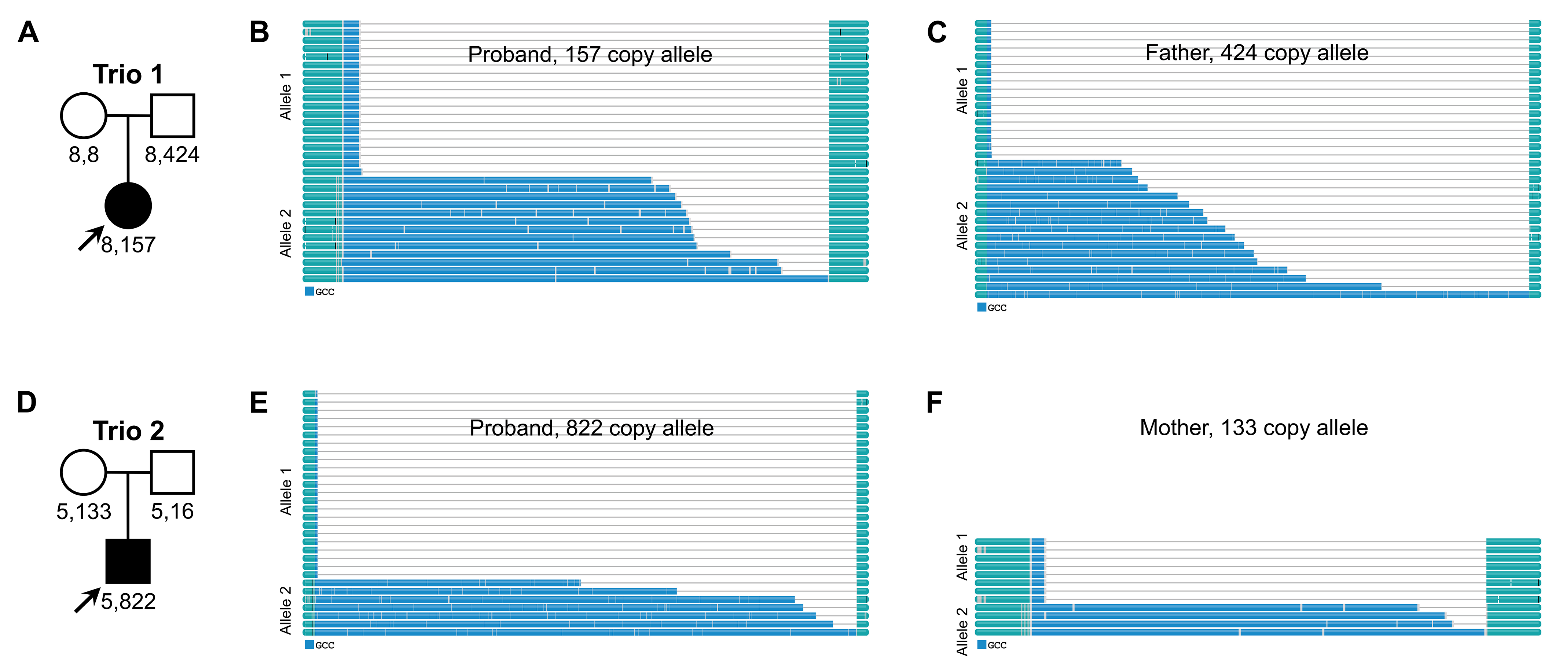

**Supplementary Figure 18.** Waterfall plots from TRGT showing length and GCC motif content of individual sequencing reads that span the *AFF3* repeat from Pacific Biosciences HiFi GS in two trios. In all four carriers of the *AFF3* expansion, the expanded allele shows significant length variation among individual reads, likely indicating somatic mosaicism for the size of the expansion. Reported size of each expanded allele is based on the consensus.

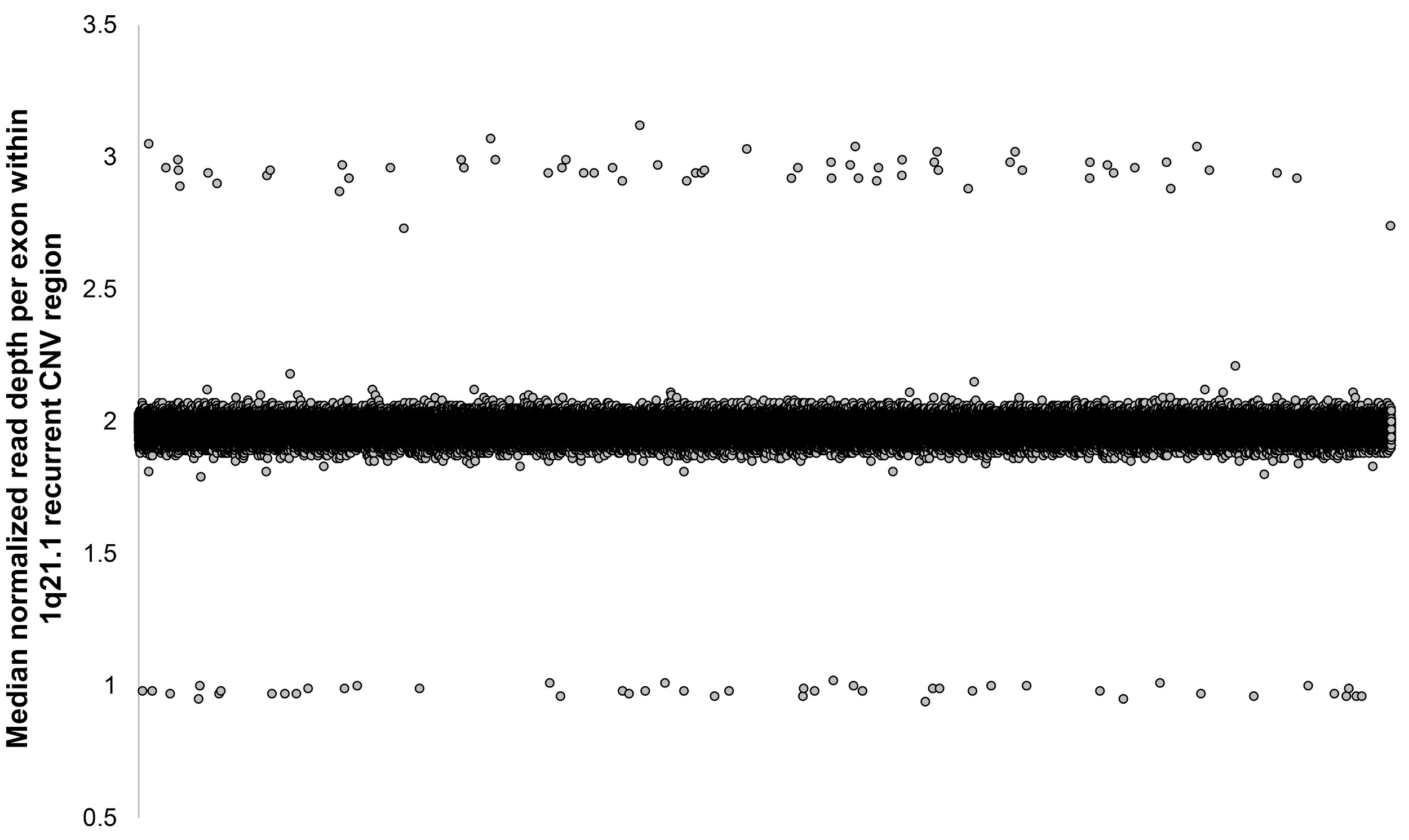

**Supplementary Figure 19. Identification of recurrent deletions and duplications in the UKB by read depth.** Each point shows the median normalized read depth per sample for all exons located within the 1q21.1 recurrent microdeletion/duplication region. Individuals with a deletion (copy number ~1) or duplication (copy number ~3) of the region are clearly visible. Similar results were obtained at the five other recurrent CNV regions analyzed.

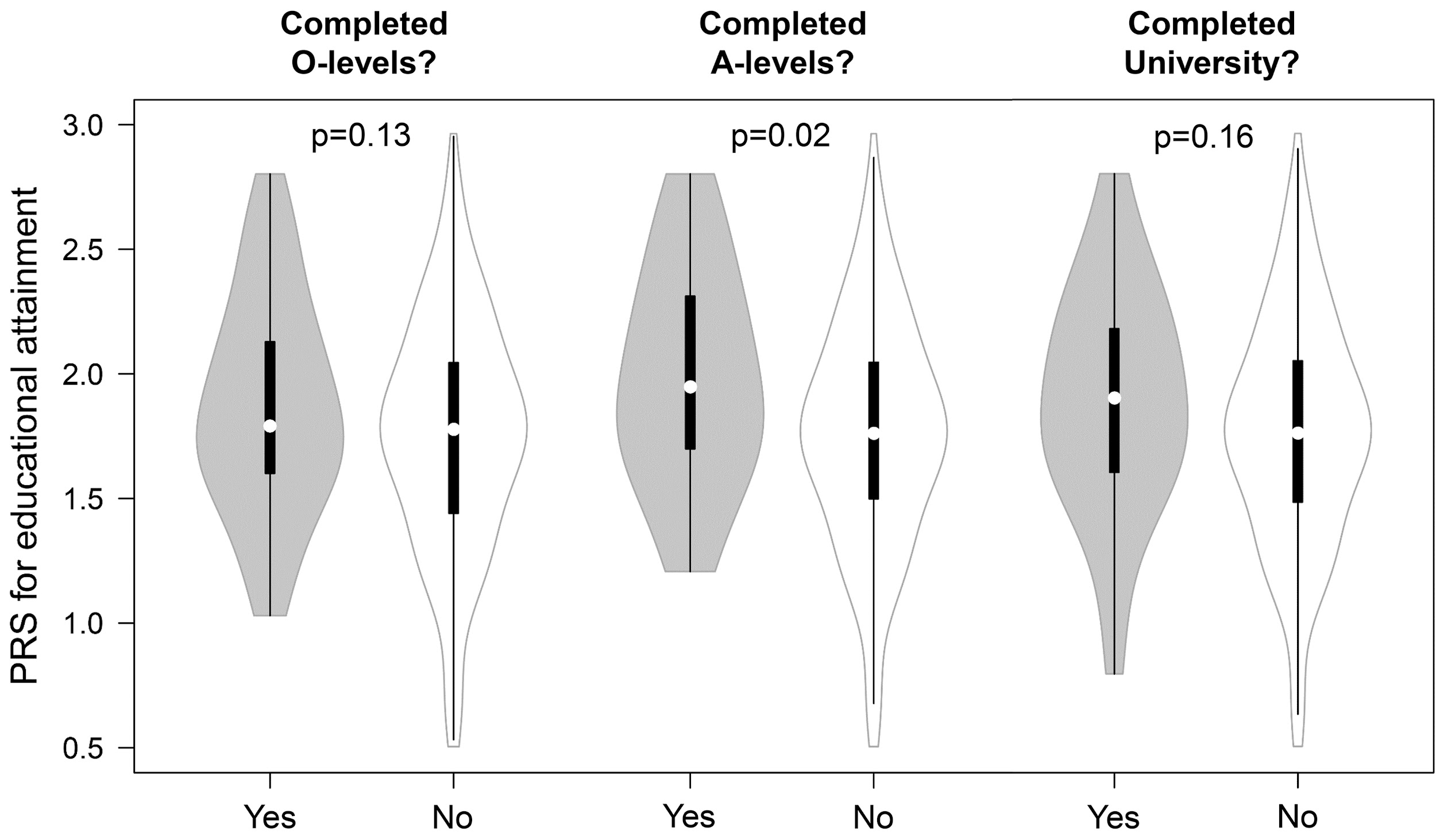

**Supplementary Figure 20. Differences in polygenic risk score underlies varying educational attainment in individuals with the *AFF3* expansion.** We divided individuals with *AFF3* expansions based on their educational attainment and compared PRS between those that did or did not attain each qualification. P-values were calculated using an unpaired Student’s t-test. Within each violin, white circles show the medians; box limits indicate the 25^th^ and 75^th^ percentiles; whiskers extend 1.5 times the interquartile range from the 25^th^ and 75^th^ percentiles.

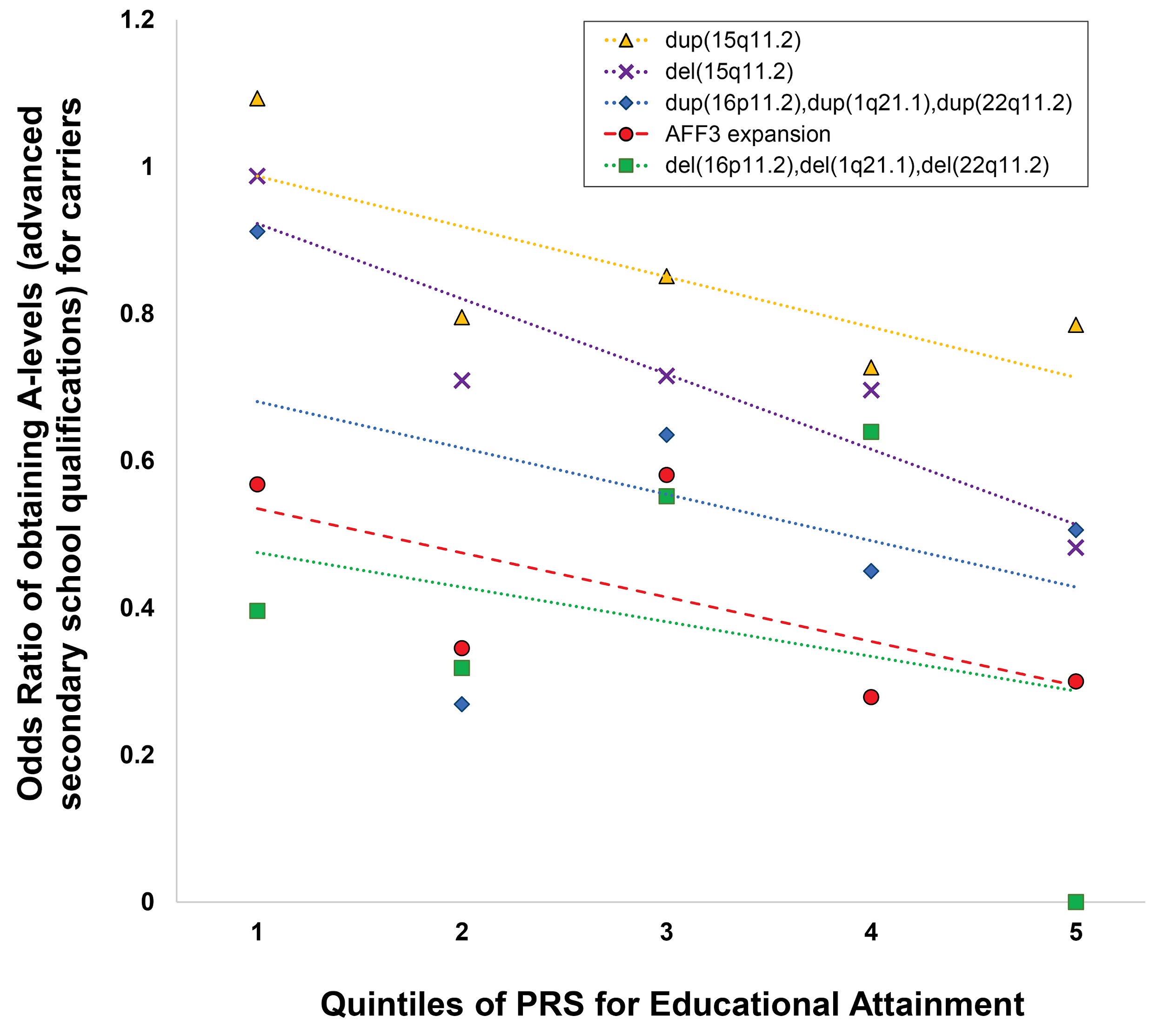

**Supplementary Figure 21. The effects of *AFF3* expansions and recurrent CNVs on educational attainment are moderated by genetic background.** Individuals were divided into quintiles based on their PRS for educational attainment and for individuals with *AFF3* expansions and recurrent pathogenic microdeletions/duplications we show the relative odds ratios of obtaining A-levels (exams taken at age 18) compared to the rest of the UKB cohort. Because of the relatively low numbers in the set of samples profiled, in order to maintain robust statistics, we grouped together individuals with either deletions or duplications of 1q21.1, 16p11.2 and 22q11.2. Quintile 1 corresponds to individuals in the top 20% of PRS for educational attainment, while quintile 5 corresponds to individuals in the lowest 20% of PRS. Dashed colored lines show the lines of best fit for each distribution.

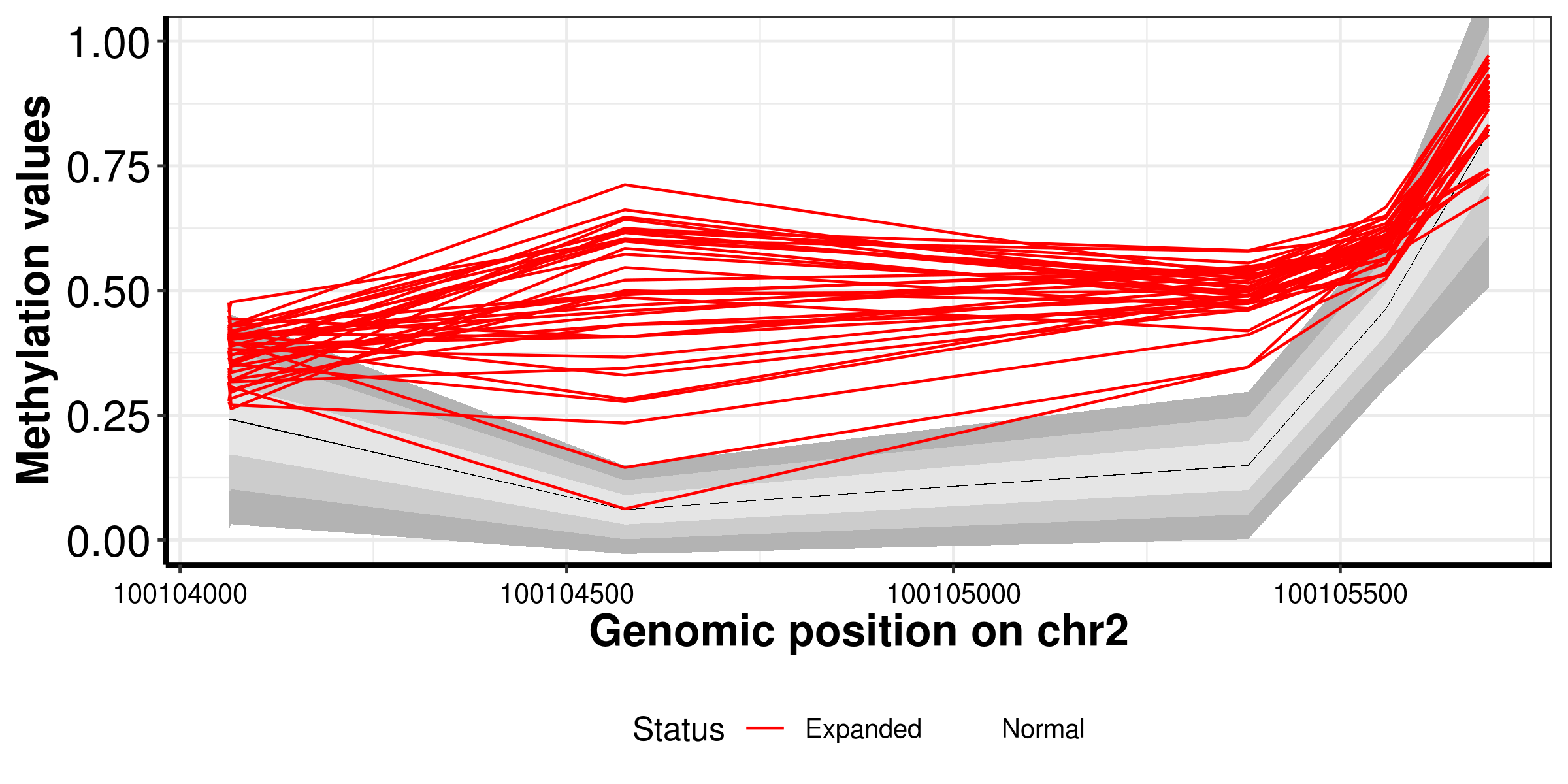

**Supplementary Figure 22.** Plot of methylation array data around the *AFF3* TR showing 38 hypermethylation events in 32,776 samples. Individuals with hypermethylation of this locus are shown as red lines. Grey shaded regions indicate 1, 1.5 and 2 standard deviations from the mean β value of the population (thin black line).

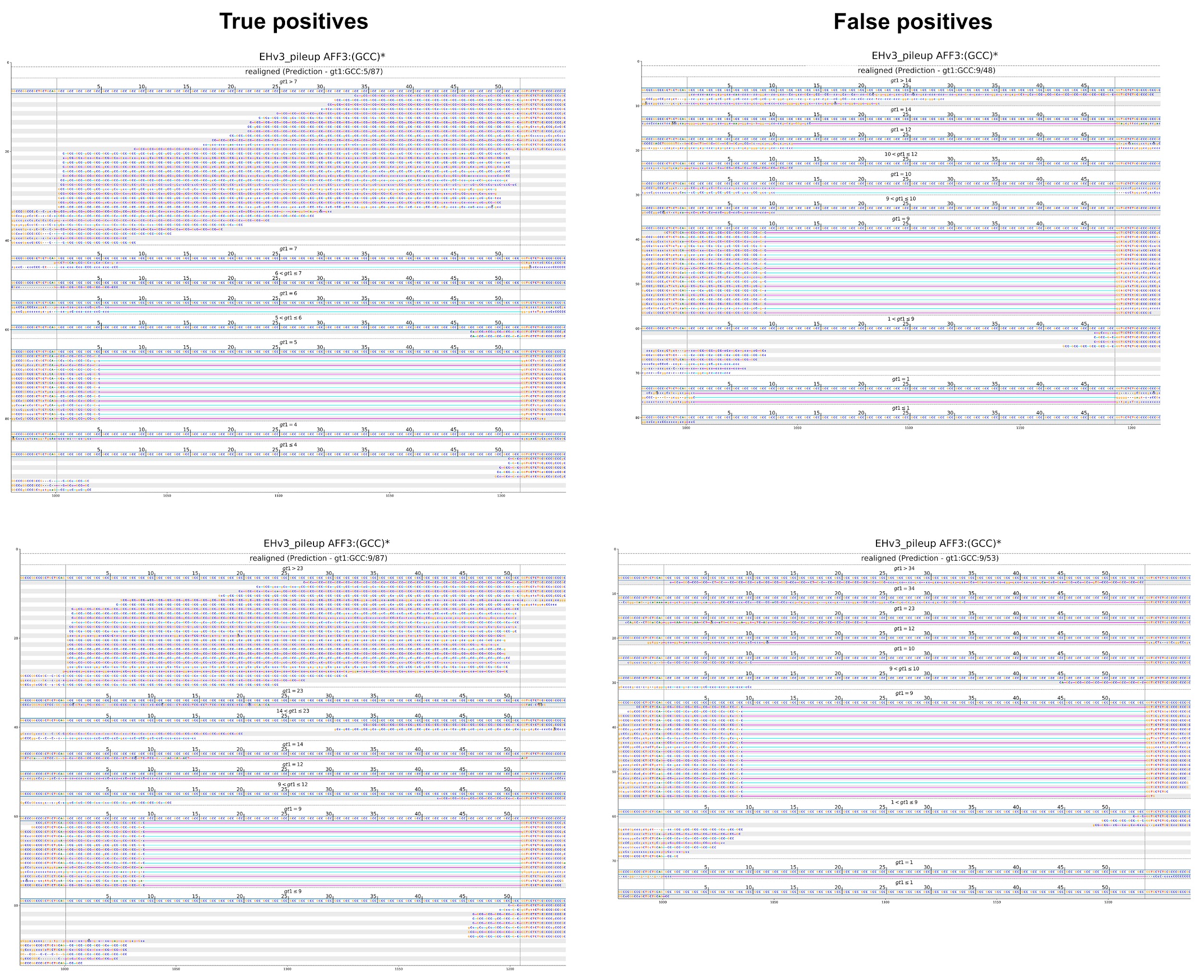

**Supplementary Figure 23. Example curation of read alignment plots in individuals identified by ExpansionHunter as having putative TREs of the GCC TR in *AFF3*.** The two examples on the left were considered as genuine TREs (true positives), as the predicted genotypes were well supported by multiple in-repeat reads. In contrast, in the two examples on the right, the putative long TR alleles identified by ExpansionHunter showed poor support, with very few reads that contain multiple mismatches aligned over the TR and were considered false positive TREs that were removed from further analysis.
